# The potential health and economic impacts of new tuberculosis vaccines under varying delivery strategies in Delhi and Gujarat, India: a modelling study

**DOI:** 10.1101/2023.09.27.23296211

**Authors:** Rebecca A Clark, Allison Portnoy, Chathika K Weerasuriya, Tom Sumner, Roel Bakker, Rebecca C Harris, Kirankumar Rade, Sanjay Kumar Mattoo, Dheeraj Tumu, Nicolas A Menzies, Richard G White

## Abstract

**Background:** India has the largest tuberculosis burden globally, but this burden varies nationwide. All-age tuberculosis prevalence in 2021 ranged from 747/100,000 in Delhi to 137/100,000 in Gujarat. Previous modelling has demonstrated the benefits and costs of introducing novel tuberculosis vaccines in India overall. However, no studies have compared the potential impact of tuberculosis vaccines in regions within India with differing tuberculosis disease and infection prevalence. We used mathematical modelling to investigate how the health and economic impact of two potential tuberculosis vaccines, M72/AS01_E_ and BCG-revaccination, could differ in Delhi and Gujarat under varying delivery strategies.

**Methods:** We applied a compartmental tuberculosis model separately for Delhi (higher disease and infection prevalence) and Gujarat (lower disease and infection prevalence), and projected epidemiological trends to 2050 assuming no new vaccine introduction. We simulated M72/AS01_E_ and BCG-revaccination scenarios varying target ages and vaccine characteristics. We estimated cumulative cases, deaths, and disability-adjusted life years averted between 2025–2050 compared to the no-new-vaccine scenario and compared incremental cost-effectiveness ratios to three cost-effectiveness thresholds.

**Results:** M72/AS01_E_ averted a higher proportion of tuberculosis cases than BCG-revaccination in both regions (Delhi: 16.0% vs 8.3%, Gujarat: 8.5% vs 5.1%) and had higher vaccination costs (Delhi: USD$118 million vs USD$27 million, Gujarat: US$366 million vs US$97 million). M72/AS01_E_ in Delhi could be cost-effective, or even cost-saving, for all modelled vaccine characteristics. M72/AS01_E_ could be cost-effective in Gujarat, unless efficacy was assumed only for those with current infection at vaccination. BCG-revaccination could be cost-effective, or cost-saving, in both regions for all modelled vaccine scenarios.

**Discussion:** M72/AS01_E_ and BCG-revaccination could be impactful and cost-effective in Delhi and Gujarat. Differences in impact, costs, and cost-effectiveness between vaccines and regions, were determined partly by differences in disease and infection prevalence, and demography. Age-specific regional estimates of infection prevalence could help to inform delivery strategies for vaccines that may only be effective in people with a particular infection status. Evidence on the mechanism of effect of M72/AS01_E_ and its effectiveness in uninfected individuals, which were important drivers of impact and cost-effectiveness, particularly in Gujarat, are also key to improve estimates of population-level impact.

## Background

India has the highest global burden of tuberculosis, but this burden varies widely across the country. In the National Tuberculosis (TB) Prevalence survey conducted from 2019–2021, the estimated tuberculosis prevalence was 312 per 100,000 for all ages in India overall.^1^ The National Capital Territory of Delhi was estimated to have the highest regional tuberculosis prevalence of 747 per 100,000, whereas Gujarat was estimated to have the lowest regional tuberculosis prevalence [137 per 100,000].^1^

Tuberculosis elimination is a key focus for the Indian government, and prevention strategies, including tuberculosis vaccines and preventive treatment, are considered within the National Strategic Plan for Elimination of Tuberculosis 2017–2025.^2^ As of July 2023, there were sixteen tuberculosis vaccine candidates in clinical trials. Results are eagerly anticipated from an upcoming Phase III trial of the vaccine candidate M72/AS01_E_ and the ongoing confirmatory Phase IIb trial for BCG-revaccination, as both products have demonstrated promising results in previous Phase IIb trials.^3,4^

Earlier modelling studies have found that the introduction of new tuberculosis vaccines could have a positive impact worldwide ^5–10^ and in India.^11–15^ However, it is unknown how or if the impact of tuberculosis vaccines will vary regionally within India, given the varying burdens of disease. The Indian government is set to undertake a study to investigate the impact of delivering BCG to household contacts aged 6–18 compared to offering preventive therapy.^16^ Variation in disease and infection prevalence may influence the impact of these interventions by region.

We used mathematical modelling to investigate how the health impact and cost-effectiveness of M72/AS01_E_ and BCG-revaccination could vary between high- and low-tuberculosis burden areas of India—represented by Delhi and Gujarat—under varying delivery strategies.

## Methods

### Data

Data to inform calibration was obtained from the National TB Prevalence survey in India,^1^ the India TB Report 2022 and 2023,^2,17^ and Ni-kshay—an online tuberculosis reporting and surveillance system developed by the National TB Elimination Programme.^18^ We combined available demographic data and extrapolated to obtain single age and year projections of population size for each region.^19^

### Model structure and calibration

We adapted a tuberculosis natural history model structure and parameterisation from previous studies.^5,11^ We employed history matching with emulation using the ‘hmer’ R package to calibrate the model to each region.^20^ We fit each model to three targets to represent the higher tuberculosis burden in Delhi, and the lower tuberculosis burden in Gujarat. We assumed a uniform distribution between lower and upper bounds, and adjusted each target as described in the Supplementary Material sections 2 and 3. We fit to the 2021 disease prevalence per 100,000 [Delhi: 747 (510–984), Gujarat: 137 (76–198)]^1^, the 2021 notification rate per 100,000 [Delhi: 536 (429–644), Gujarat: 137 (110–165)]^17^, and the 2020 proportion of active tuberculosis that was subclinical [0.564 (interquartile range = 0.428–0.685)]^21^. The model for Gujarat was also fit to the estimated adult tuberculosis prevalence in 2011 [383 (315–451) per 100,000].^22^

### Scenarios

#### i. No-new-vaccine baseline

We used the calibrated models for Delhi and Gujarat to project baseline epidemiology to 2050 in each setting, assuming the coverage and quality of non-vaccine tuberculosis services continued at 2019 levels, with no new vaccine introduction.

#### ii. Vaccine scenarios

We established a *Basecase* vaccine scenario for each vaccine product. *Basecase* vaccine characteristics were informed by trial characteristics and expert opinion, and we assumed that each vaccine would be delivered to an age group aligned with the clinical-trial-eligible ages.^3,4^ The *Basecase* M72/AS01_E_ scenario assumed a 50% efficacy prevention of disease vaccine effective with any infection status at vaccination and ten years average protection, introduced in 2030 routinely to those aged 15 (achieving 80% coverage over five years) and as a campaign for ages 16–34 (achieving 70% coverage over five years) in 2030 and 2040. The *Basecase* BCG-revaccination scenario assumed a 45% efficacy prevention of infection vaccine effective in individuals with no current infection at the time of vaccination and ten years average protection, introduced in 2025 routinely to those aged 10 (achieving 80% coverage over five years) and as a campaign for ages 11–18 (achieving 80% coverage over five years), in 2025, 2035, and 2045.

We evaluated age-targeting *Policy Scenarios* for both vaccine products. We met with in-country partners in the Government of India to discuss preferred ages to target for tuberculosis vaccine delivery. We ensured that our modelled scenarios captured this information to provide the most useful estimates to decision makers. The *Older Ages: M72/AS01_E_* scenario assumed routine delivery to those aged 17 and a campaign for ages 18–55, and the *Older Ages: BCG-revaccination* scenario assumed routine delivery to those aged 15 and a campaign for ages 16–34. For both vaccine products, we evaluated an *All-Adults* scenario with routine delivery for those aged 18 and a campaign for everyone aged 19 and older.

To investigate uncertainty in vaccine product characteristics, we evaluated *Vaccine Characteristic and Coverage Scenarios* by varying individual features of the vaccine profile from the *Basecase* (Table 1).

**Table 1.** Vaccine scenarios.

| Characteristic | M72/AS01<sub>E</sub> |  | BCG-revaccination |  |
| --- | --- | --- | --- | --- |
|  | Basecase | Univariate scenario analyses | Basecase | Univariate scenario analyses |
| *Policy Scenarios* |  |  |  |  |
| Age targeting | Routine for age 15, campaign for ages 16–34 | *Older Ages* (routine for age 17, campaign for ages 18–55)<br>*All-Adults* (routine for age 18, campaign for ages 19+) | Routine for age 10, campaign for ages 11–18 | *Older Ages* (routine for age 15, campaign for ages 16–34)<br>*All-Adults* (routine for age 18, campaign for ages 19+) |
| *Vaccine Characteristic and Coverage Scenarios* |  |  |  |  |
| Efficacy | 50% | 60%, 70% | 45% | 70% |
| Mechanism of effect | Prevents disease | Prevents infection and disease | Prevents infection | Prevents infection and disease |
| Infection status at time of vaccination required for efficacy | Any infection (current / no current infection) | Current infection only | No current infection only | Any infection (current / no current infection) |
| Duration of protection | 10 years | 5, 15, 20 | 10 years | 5, 15, 20 |
| Introduction year | 2030 | 2036 | 2025 | 2031 |
| Coverage | Medium:<br>80% routine, 70% campaign | Low:<br>70% age 15, 50% campaign<br>High:<br>90% age 15, 90% campaign | Medium:<br>80% routine, 80% campaign | Low:<br>70% routine, 70% campaign<br>High:<br>90% routine, 90% campaign |

We assumed vaccine delivery costs of $2.50 (1.00–5.00) per dose, supply chain costs of $0.11 (0.06–0.22) per dose and a vaccine price of $2.50 per dose for M72/AS01_E_ (assuming two doses per course) and $0.17 per dose for BCG-revaccination (assuming one dose per course). For vaccine campaigns, we included a one-time vaccine introduction cost of $2.40 (1.20–4.80) per individual in the targeted age group to represent non-recurring start-up costs.

### Outcomes

We estimated the cumulative number of tuberculosis cases and deaths averted between vaccine introduction and 2050 for each scenario compared to the predicted numbers in the no-new-vaccine baseline. We estimated incidence and mortality rate reductions in 2050 for each scenario compared to the estimated rates in 2050 for the no-new-vaccine baseline. We calculated incremental vaccination, diagnostic, and treatment costs for each scenario compared to the no-new-vaccine baseline in 2020 US dollars.

We performed cost-effectiveness analysis comparing the *Policy Scenarios* for each vaccine product and region. Costs and benefits were discounted to 2025 at 3% per year as per guidelines.^23^ We estimated incremental costs and disability-adjusted life years (DALYs) averted for each scenario between 2025–2050, using the disability weight for tuberculosis from the Global Burden of Disease 2019 study,^24^ and India-specific life expectancy estimates from the United Nations Development Programme.^25^ We calculated incremental cost-effectiveness ratios (ICERs) as mean incremental costs divided by mean incremental DALYs averted for each scenario. We evaluated the resulting ICERs against three cost-effectiveness thresholds: 1 times gross domestic product (GDP) per capita for India (US$1,928), and two opportunity cost thresholds defined by Ochalek et al: the country-level upper (US$443) and lower (US$328) bounds.^26^

To investigate if the decision to introduce a vaccine would change based on the assumed vaccine characteristics, we calculated ICERs for the *Vaccine Characteristic and Coverage Scenarios* compared to the no-new-vaccine baseline. We assumed each vaccine product was delivered using the *Basecase* age-targeting assumptions.

## Results

Calibrated trends for Delhi and Gujarat are shown in Figure 1. Between 2025 and 2050, the no-new-vaccine baseline predicted 4.1m (95% uncertainty interval: 3.7–4.4) cases and 533 (349–761) thousand deaths in Delhi, and 2.2m (2.0–2.5) cases and 210 (100–325) thousand deaths in Gujarat. Consistent with findings from the National TB Prevalence Survey, a higher burden of disease was predicted in Delhi than in Gujarat. A lower and declining trend in tuberculosis infection prevalence was predicted in Gujarat compared to Delhi.

**Figure 1.**
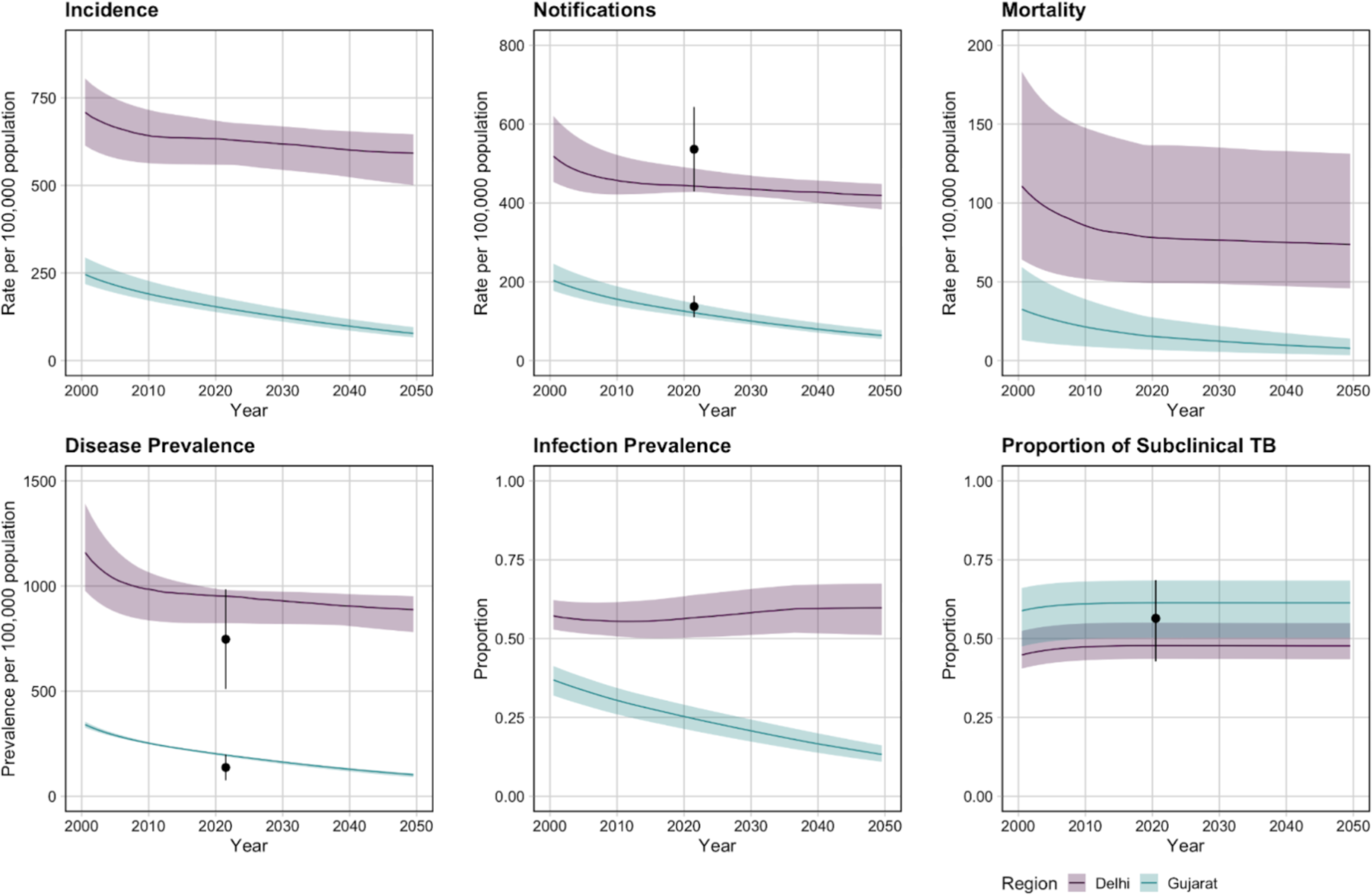
Calibrated epidemiological trends for Delhi and Gujarat

Key results are described below, with full results in Supplementary Material sections 8 and 9. The *Basecase* M72/AS01_E_ scenario averted 655 (587–730) thousand cases, or 16.0% of the total predicted cases, and 77 (49–112) thousand deaths, or 14.4% of the total predicted deaths between 2025 and 2050 in Delhi (Table 2). The *Basecase* M72/AS01_E_ scenario averted 186 (155–228) thousand cases (8.5% of the total predicted cases) and 16 (7–27) thousand deaths (7.6% of the total predicted deaths) in Gujarat between 2025–2050 (Table 2). The number of cases and deaths averted was increased in both Delhi and Gujarat with delivery to an older population (Table 2). The *All-Adults* scenario averted more cases and deaths than the *Older Ages* scenario, which similarly averted more than the *Basecase* M72/AS01_E_ scenario (Figure 2).

**Table 2.** Health impact results for M72/AS01_E_ and BCG-revaccination in Delhi and Gujarat.

| Scenario | Cumulative cases averted between 2025–2050 (1000s) |  | Cumulative deaths averted between 2025–2050 (1000s) |  | Incidence rate reduction in 2050 (%) |  | Mortality rate reduction in 2050 (%) |  |
| --- | --- | --- | --- | --- | --- | --- | --- | --- |
|  | Delhi | Gujarat | Delhi | Gujarat | Delhi | Gujarat | Delhi | Gujarat |
| **M72/AS01<sub>E</sub> scenarios** |  |  |  |  |  |  |  |  |
| Basecase<br>(routine age 15, campaign ages 16–34) | 655<br>(587–730) | 186<br>(155–228) | 77<br>(49–112) | 16<br>(7–27) | 26<br>(23–29) | 16<br>(15–19) | 27<br>(23–30) | 17<br>(15–19) |
| *Policy Scenarios* |  |  |  |  |  |  |  |  |
| Older ages<br>(routine age 17, campaign ages 18–55) | 839<br>(755–932) | 331<br>(284–393) | 98<br>(63–143) | 28<br>(13–46) | 29<br>(25–33) | 25<br>(23–27) | 31<br>(26–34) | 26<br>(24–28) |
| All-adults<br>(routine age 18, campaign ages 19+) | 935<br>(836–1,037) | 492<br>(434–575) | 108<br>(70–157) | 42<br>(20–66) | 31<br>(26–34) | 32<br>(30–34) | 32<br>(27–36) | 34<br>(32–36) |
| *Vaccine Characteristic and Coverage Scenarios* |  |  |  |  |  |  |  |  |
| Efficacy with current infection at vaccination | 471<br>(403–535) | 101<br>(84–124) | 55<br>(34–82) | 9<br>(4–15) | 17<br>(16–19) | 8<br>(7–9) | 18<br>(16–20) | 8<br>(7–9) |
| Prevention of infection and disease | 817<br>(730–914) | 238<br>(198–293) | 95<br>(61–140) | 20<br>(9–34) | 33<br>(29–37) | 22<br>(19–25) | 34<br>(29–38) | 22<br>(20–25) |
| **BCG-revaccination scenarios** |  |  |  |  |  |  |  |  |
| Basecase<br>(routine age 10, campaign ages 11–18) | 359<br>(305–402) | 113<br>(92–143) | 44<br>(29–65) | 10<br>(5–17) | 13<br>(10–16) | 10<br>(9–12) | 14<br>(10–16) | 10<br>(9–12) |
| *Policy Scenarios* |  |  |  |  |  |  |  |  |
| Older ages<br>(routine age 15, campaign ages 16–34) | 287<br>(196–352) | 152<br>(125–188) | 33<br>(20–51) | 13<br>(6–22) | 10<br>(6–14) | 13<br>(11–15) | 10<br>(6–14) | 9<br>(8–11) |
| All-adults<br>(routine age 18, campaign ages 19+) | 224<br>(139–287) | 184<br>(155–222) | 25<br>(15–40) | 16<br>(7–26) | 8<br>(4–11) | 15<br>(13–17) | 8<br>(4–11) | 11<br>(10–13) |
| *Vaccine Characteristic and Coverage Scenarios* |  |  |  |  |  |  |  |  |
| Efficacy with any infection at vaccination | 434<br>(390–494) | 114<br>(92–145) | 54<br>(34–80) | 10<br>(5–17) | 16<br>(13–19) | 10<br>(9–12) | 17<br>(14–19) | 10<br>(9–12) |
| Prevention of infection and disease | 544<br>(490–601) | 154<br>(125–195) | 67<br>(43–98) | 14<br>(6–23) | 21<br>(17–24) | 14<br>(12–16) | 21<br>(18–24) | 13<br>(12–16) |

*Estimates are provided as the median and 95% uncertainty intervals*

**Figure 2.**
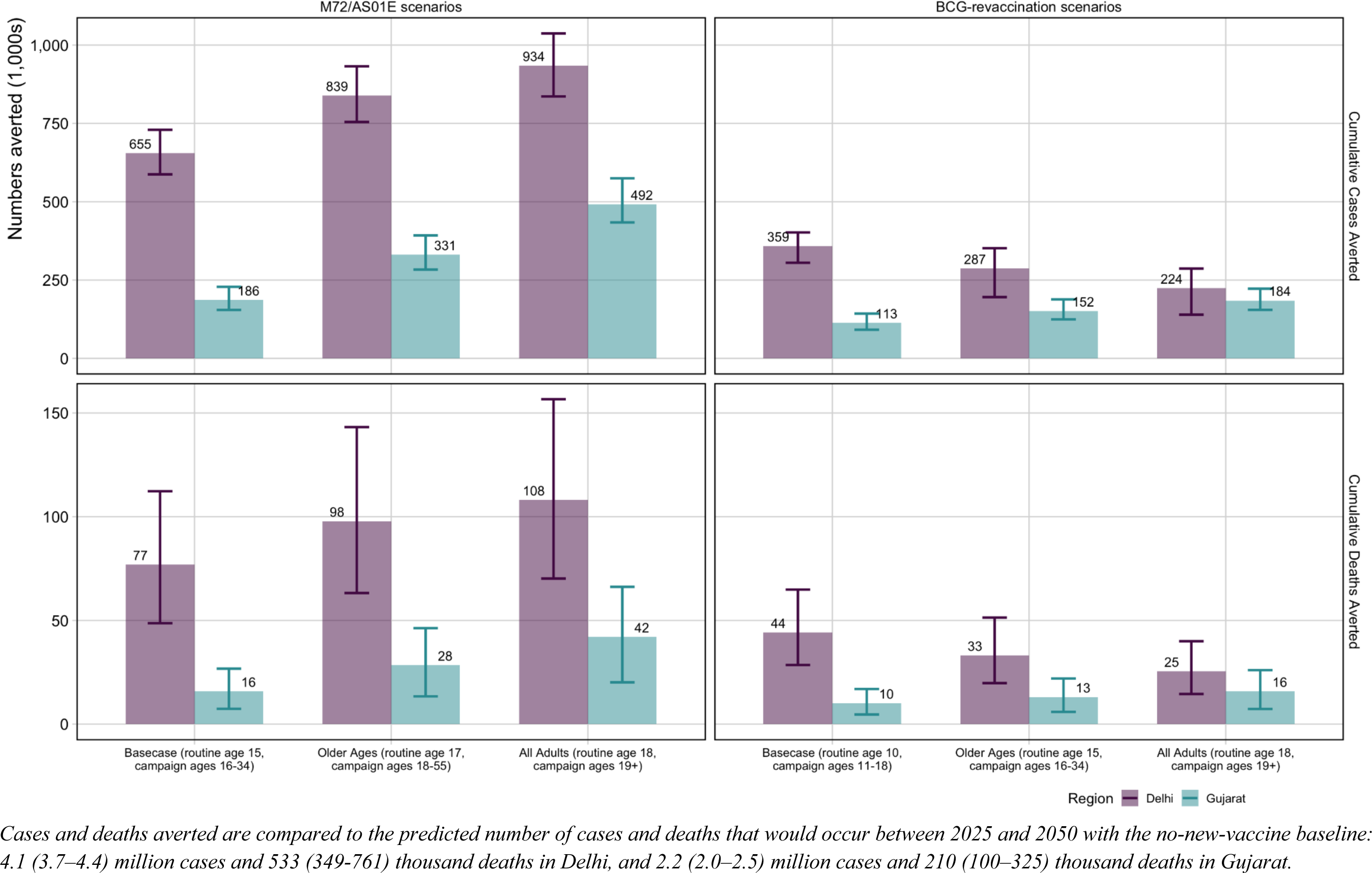
Cumulative cases and deaths averted between 2025–2050 for *Policy Scenarios* for both vaccines and regions

If M72/AS01_E_ was able to prevent both infection and disease, the number of cases and deaths averted could increase by 23–25% in Delhi and 25–28% in Gujarat compared to the *Basecase* M72/AS01_E_ scenario (Table 2). However, if M72/AS01_E_ was only efficacious with current infection at vaccination, the number of cases and deaths averted could decrease by 28–29% in Delhi and 44–46% in Gujarat compared to the *Basecase* M72/AS01_E_ scenario (Table 2).

The *Basecase* BCG-revaccination scenario averted 359 (305–402) thousand cases (8.8% of total predicted cases) and 44 (29–65) thousand deaths (8.3% of total predicted deaths) in Delhi, and 113 (91–143) thousand cases (5.1% of total predicted cases) and 10 (5–17) thousand deaths (4.8% of total predicted deaths) in Gujarat between 2025–2050 (Table 2). Due to differences in modelled infection prevalence, delivering BCG-revaccination to an older population (*Older Ages* and *All-Adults* scenarios) decreased the number of cases and deaths averted in Delhi, but increased the impact in Gujarat compared to the *Basecase* BCG-revaccination scenario (Figure 2).

If BCG-revaccination was able to prevent infection and disease, the absolute number of cases and deaths averted could increase by 52–53% in Delhi and 36–40% in Gujarat compared to the *Basecase* BCG-revaccination scenario (Table 2). If BCG-revaccination worked in any infection status opposed to only those who were uninfected, the number of cases and deaths averted could increase by 21–23% in Delhi, but could only increase the number of cases and deaths averted in Gujarat by 0–1% compared to the *Basecase* BCG-revaccination scenario (Table 2).

In both regions, M72/AS01_E_ resulted in a higher number of cases and deaths averted than BCG-revaccination: approximately 1.8 times in Delhi and 1.6 times in Gujarat (Table 2). For both vaccine products, more cases and deaths were averted in Delhi compared to Gujarat: 3.5–4.8 times for M72/AS01_E_ and 3.2–4.4 times for BCG-revaccination (Table 2).

The total vaccination cost for the M72/AS01_E_ *Basecase* was US$118m (80–173) in Delhi and was US$366m (248–536) in Gujarat, compared to the BCG-revaccination Basecase vaccination total cost of US$27m (12–49) in Delhi and US$97m (42–178) in Gujarat (Tables S10.2, S10.5, S10.8, S10.11). Larger vaccination costs were predicted for introducing M72/AS01_E_ compared to BCG-revaccination in both regions: 4.4 times more in Delhi and 3.8 times more in Gujarat. Incorporating cost-savings in treatment and diagnostic costs, the total incremental programme cost for the M72/AS01_E_ *Basecase* in Delhi was US$5m (minus 37–63) and in Gujarat was US$332m (213– 505) (Tables S10.2, S10.8). The *Basecase* BCG-revaccination scenario led to cost-savings of US$38m (58–13) in Delhi (Table S10.5). The total programme cost for the *Basecase* BCG-revaccination scenario in Gujarat was US$77m (21–158) in Gujarat (Table S10.11).

In Delhi, introducing M72/AS01_E_ was potentially cost-effective for all *Policy Scenarios*. The *Basecase* M72/AS01_E_ scenario (ICER = US$4), *Older Ages* scenario (ICER = US$126) and *All-Adults* scenario (ICER = US$317) were cost-effective at the country-level upper and lower bounds, and the 1xGDP threshold (Table 3, Figure 3). The incremental cost of the *Basecase* M72/AS01_E_ scenario was US$5m (minus 37–63), averting 1.5m (1.0–2.1) DALYs between 2025–2050 compared to the no-new-vaccine baseline (Table 3, Figure 3). In Gujarat, only the *All-Adults* scenario was considered potentially cost-effective for M72/AS01_E_ at the 1xGDP threshold (ICER = US$975) (Table 3, Figure 3). The cost of the *All-Adults* scenario compared to the no-new-vaccine baseline was US$624m and 640 thousand DALYs were averted between 2025–2050 (Table 3, Figure 3).

**Table 3.** Competing choice cost-effectiveness analysis for Delhi and Gujarat.

| Scenario | Total costs<br>(USD, 1000s) | Total DALYs<br>averted<br>(1000s) | Incremental cost<br>(USD, 1000s) | Incremental<br>DALYs averted<br>(1000s) | Cost (USD) per<br>DALY averted |
| --- | --- | --- | --- | --- | --- |
| **Delhi** |  |  |  |  |  |
| **M72/AS01<sub>E</sub> policy scenarios** |  |  |  |  |  |
| No-new-vaccine | 977,788 | – | – | – | – |
| Basecase (routine age 15,<br>campaign for ages 16–34) | 982,966 | 1,465 | 5,178 | 1,465 | 4 |
| Older ages (routine age 17,<br>campaign for ages 18–55) | 1,023,279 | 1,786 | 40,313 | 321 | 126 |
| All-adults (routine age 18,<br>campaign for ages 19+) | 1,050,875 | 1,873 | 27,596 | 87 | 317 |
| **BCG-revaccination policy scenarios** |  |  |  |  |  |
| No-new-vaccine | 977,788 | – | – | – | – |
| Basecase (routine age 10,<br>campaign for ages 11–18) | 940,220 | 938 | -37,568 | 938 | *Cost-saving* |
| Older ages (routine age 15,<br>campaign for ages 16–34) | 973,930 | 693 | – | – | *Strongly*<br>*dominated* |
| All-Adults (routine age 18,<br>campaign for ages 19+) | 1,032,616 | 521 | – | – | *Strongly*<br>*dominated* |
| **Gujarat** |  |  |  |  |  |
| **M72/AS01<sub>E</sub> policy scenarios** |  |  |  |  |  |
| No-new-vaccine | 584,609 | – | – | – | – |
| Basecase (routine age 15,<br>campaign for ages 16–34) | 917,077 | 308 | – | – | *Weakly*<br>*dominated* |
| Older ages (routine age 17<br>campaign for ages 18–55) | 1,097,770 | 505 | – | – | *Weakly*<br>*dominated* |
| All-Adults (routine age 18,<br>campaign for ages 19+) | 1,208,573 | 640 | 623,965 | 640 | 975 |
| **BCG-revaccination policy scenarios** |  |  |  |  |  |
| No-New-Vaccine | 584,609 | – | – | – | – |
| Basecase (routine age 10,<br>campaign for ages 11-18) | 661,265 | 219 | 76,656 | 219 | 351 |
| Older ages (routine age<br>15, campaign ages 16–34) | 708,672 | 273 | 47,407 | 55 | 868 |
| All-Adults (routine age<br>18, campaign ages 19+) | 844,338 | 312 | 135,666 | 39 | 3,486 |

**Figure 3.**
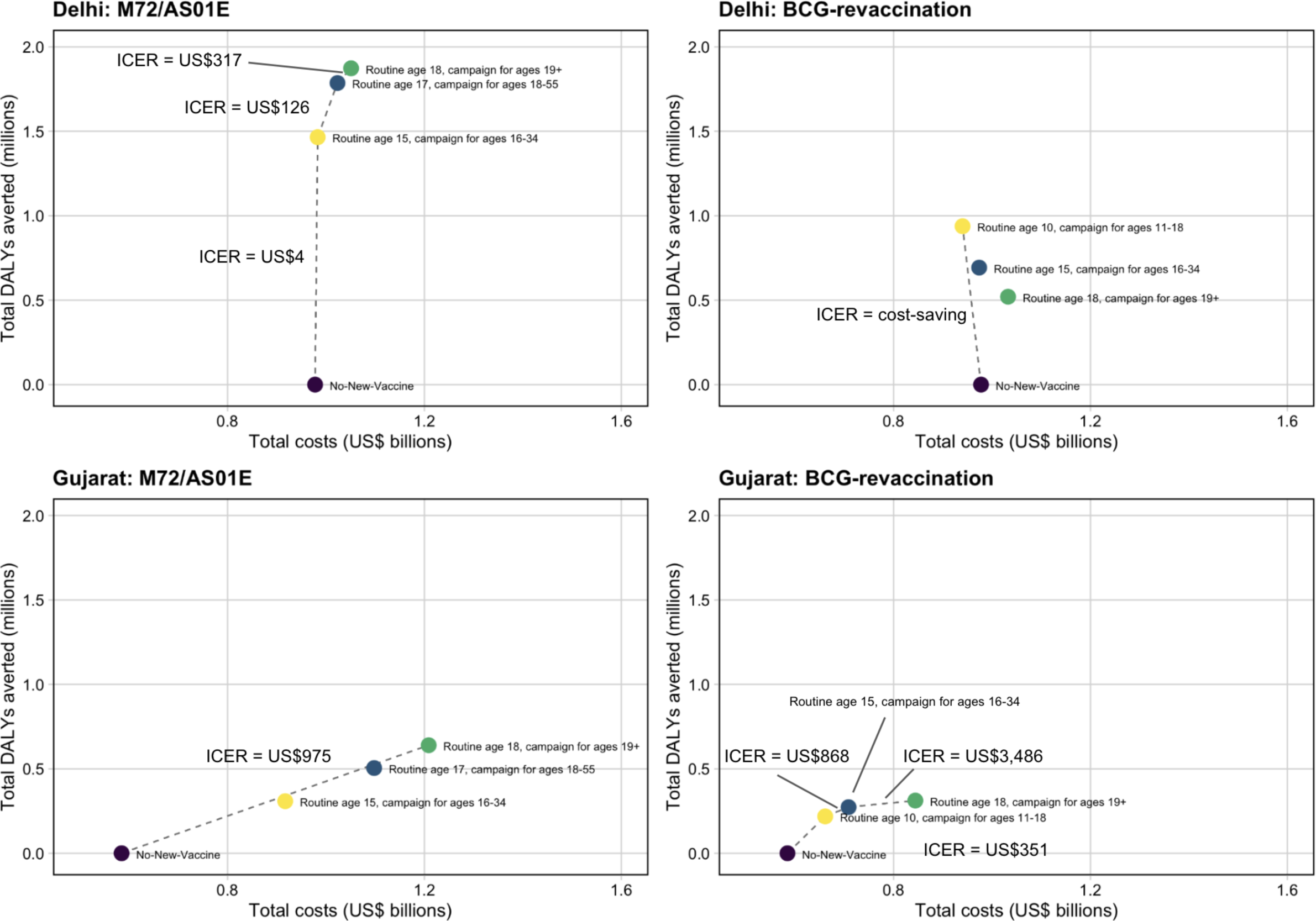
Competing choice cost-effectiveness analysis for Delhi and Gujarat *Policy Scenarios* for both vaccine products

In Delhi, the *Older Ages* and *All-Adults* BCG-revaccination scenarios were dominated by the *Basecase* BCG-revaccination scenario. The *Basecase* BCG-revaccination scenario was considered cost-effective at all thresholds (ICER = cost-saving), with cost-savings of US$37m and averted 938 thousand DALYs between 2025–2050 compared to the no-new-vaccine baseline. In Gujarat, the *Basecase* BCG-revaccination scenario was cost-effective at the country-level upper bound (ICER = US$351), with an incremental cost of US$77m compared to the no-new-vaccine baseline and averted 219 thousand DALYs between 2025–2050. The *Older Ages* scenario was cost-effective at 1xGDP per capita (ICER = US$868) (Table 3, Figure 3).

When comparing the ICERs from the *Vaccine Characteristic and Coverage Scenarios* in Delhi, regardless of the assumed product characteristics, introducing M72/AS01_E_ routinely to those aged 15 and as a campaign for ages 16–34 could be cost-effective, and in some cases, cost-saving, at the country-level lower bound (Figure 4). Similarly, introducing BCG-revaccination routinely to those aged 10 and as a campaign for ages 11–18 could be cost-saving in Delhi (Figure 4). In Gujarat, delivering M72/AS01_E_ routinely to those aged 15 and as a campaign for ages 16–34 could be cost-effective at a 1xGDP per capita threshold, except if the vaccine was only efficacious with current infection at vaccination (Figure 4). Introducing BCG-revaccination in Gujarat could be cost-effective regardless of the assumed product characteristics (Figure 4).

**Figure 4.**
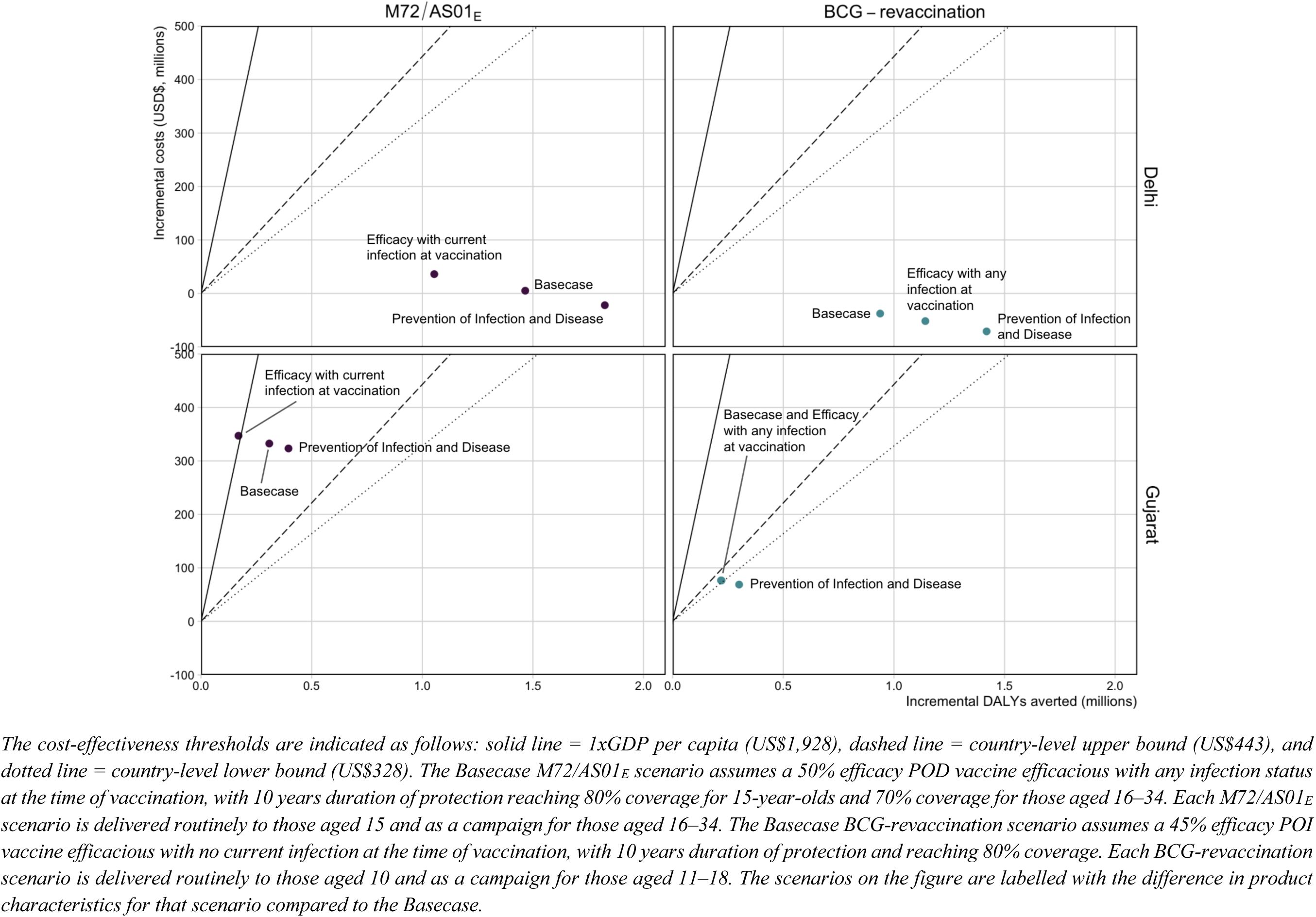
Comparison of ICERs for select *Vaccine Characteristic and Coverage Scenarios*

In both regions, there were larger ICERs for M72/AS01_E_ scenarios compared to BCG-revaccination, and for both vaccine products, larger ICERs for Gujarat compared to Delhi (Figure 4). Full impact results are in Supplementary Material sections 8 and 9.

## Discussion

Our modelling suggests that M72/AS01_E_ and BCG-revaccination could have a substantial impact in Delhi and Gujarat. M72/AS01_E_ scenarios resulted in a higher number of cases and deaths averted than BCG-revaccination in both regions, and more cases and deaths were averted in Delhi compared to Gujarat. We found that given the assumed characteristics, both products were likely to be cost-effective or cost-saving in Delhi. In Gujarat, M72/AS01_E_ was likely to be cost-effective unless it only worked in those with current infection at the time of vaccination. M72/AS01_E_ scenarios had higher vaccination costs than BCG-revaccination, and higher vaccination costs were estimated in Gujarat overall than in Delhi.

For all modelled scenarios, M72/AS01_E_ would have a larger and faster impact on the tuberculosis burden than BCG-revaccination. We assumed that M72/AS01_E_ would be effective regardless of the presence or absence of infection, and work by preventing disease. Therefore, those with current infection who received the vaccine would have an immediately lower rate of disease progression. We assumed that BCG-revaccination would only be effective in those who were uninfected at vaccination and would work by preventing infection. Therefore, the impact from BCG-revaccination would be delayed by the typical time from vaccination to infection, and the typical time from infection to disease.

Several findings related to the lower infection prevalence modelled in Gujarat compared to Delhi. For M72/AS01_E_ scenarios, the relative decrease in the number of cases and deaths averted if M72/AS01_E_ was only effective in individuals with current infection was much larger in Gujarat compared to Delhi. If M72/AS01_E_ vaccine efficacy was restricted to those with current infection, a larger proportion of the population would no longer benefit from vaccination in Gujarat compared to Delhi, due to the lower infection prevalence in Gujarat. BCG-revaccination was estimated to have a larger relative impact in Gujarat than in Delhi for strategies targeting an older and larger proportion of the population (*Older Ages* or *All-Adults* scenarios compared to the *Basecase*). As we modelled a higher infection prevalence for all ages in Delhi, and assumed that BCG-revaccination would only be effective if administered to people who were uninfected, there was a higher proportion of the population who were uninfected and would receive protection from the vaccine in Gujarat than in Delhi.

Across the range of assumptions examined for vaccine product characteristics, M72/AS01_E_ and BCG-revaccination were likely to be cost-effective (and even cost-saving) in Delhi compared to the thresholds evaluated. In Gujarat, M72/AS01_E_ could be cost-effective unless efficacy was restricted to those with current infection, and BCG-revaccination was likely to be cost-effective regardless of the modelled characteristics. Understanding the mechanism of effect of M72/AS01_E_, and confirming whether it works in all populations is a key area for future research, particularly in Gujarat and other areas with a low prevalence of infection.

M72/AS01_E_ was predicted to have higher vaccination costs than BCG-revaccination in both regions: 4.4 times as high in Delhi (US$118m vs US$27m) and 3.5 times as high in Gujarat (US$366m vs US$97m), due to the higher price per dose for M72/AS01_E_, ($2.50 per dose vs $0.17 per dose for BCG) and assuming two doses per course. Higher costs for both products were predicted in Gujarat compared to Delhi due to the larger population size.

There are limitations associated with this work. Firstly, this is a mathematical modelling study, and therefore limitations associated with models apply. We represented tuberculosis natural history with a compartmental model accounting for multiple infection states. If our assumptions around how the latency structure or aspects such as subclinical tuberculosis interact with vaccines were incorrect, we may have over- or under-protected the population, leading to incorrect impact estimates. We assumed bounds of certain natural history parameters would not vary between regions in India, and therefore used national India posterior ranges as priors for Delhi and Gujarat calibration.^11^ If this was an incorrect assumption, or if initial assumptions on the national India model prior ranges were incorrect, our projections may inaccurately represent Delhi and Gujarat.

Our model included an on-treatment compartment but assumed the only people treated were those with disease. The reported notification rate in Gujarat was greater than the prevalence estimate, implying more people were treated per year than those with prevalent disease. While Gujarat has excellent tuberculosis treatment services, only 35% of reported notifications in 2021 were bacteriologically confirmed. Therefore, there could be treatment of individuals who did not have tuberculosis, which we did not represent, but could be investigated with future adaptations to the model.

A key limitation of this work was the availability of region-specific data to inform calibration. The National TB Prevalence Survey in India provided estimates of the tuberculosis prevalence for each region for one year, allowing us to model a higher burden of tuberculosis in Delhi compared to Gujarat, but this did not allow us to incorporate a data-driven time trend. There were no region-specific calibration targets to constrain mortality, and therefore we found large uncertainty on the number of cumulative deaths averted due to large uncertainty around trends in mortality. Additionally, there were no region-specific estimates of infection prevalence, which was a key determiner of vaccine impact. We assumed that differences in mortality and infection prevalence between Delhi and Gujarat would align with the differences observed in disease prevalence, and modelled a higher mortality rate and infection prevalence in Delhi. To continue modelling subnational regions, more region-specific data to inform model predictions is urgently needed.

We represented population size and age structure for Delhi and Gujarat by utilising all available demographic data and projections for the regions and extrapolated forward to 2050 where no data was available. As the risk of tuberculosis is age-dependent, if we incorrectly represented the demographic structure of the regions we may have over or underestimated the health impact possible with new vaccines.

The no-new-vaccine baseline assumed that the current quality and coverage of services would continue. We did not consider improvements in social determinants which may occur over the time-period. If the burden projected in the no-new-vaccine baseline was higher than reality, we may be overestimating the health benefit and cost-effectiveness of vaccines. We introduced vaccines into the population independently, and did not integrate with other available services, such as tuberculosis preventive therapy, which may alter future outcomes.

## Conclusions

Our study has demonstrated that M72/AS01_E_ and BCG-revaccination are likely to be impactful and cost-effective if introduced in Delhi and Gujarat. Delhi and Gujarat were selected as the modelled regions to represent a high and low burden setting respectively. There were differences in vaccine impact between regions, which were only revealed through subnational modelling and considering differences in disease and infection prevalence. While national models are beneficial to demonstrate potential impact overall, if there are distinct epidemiological differences within the country the impact can vary.

Our results support the need for more infection prevalence surveys. We discovered how important the modelled infection prevalence of each region was to determine the likely impact if vaccines may only work in those who are uninfected or those who are infected. Age-specific regional estimates of infection prevalence would help to inform delivery strategies for vaccines only effective in people with a particular infection status, and improve estimates of vaccine impact. Another key area for future research is investigating the mechanism of effect of M72/AS01_E_, and confirming effectiveness in uninfected individuals, which was an important driver of impact and cost-effectiveness in Gujarat. Further research to reduce vaccine characteristic uncertainty and generate subnational models for additional regions is needed to maximise success of vaccine delivery in India.

## Data sharing statement

No individual level participant data was used for this modelling study. Analytic code will be made available at https://doi.org/10.5281/zenodo.6421372 immediately following publication indefinitely for anyone who wishes to access the data for any purpose.

## Data Availability

Analytic code will be made available at https://doi.org/10.5281/zenodo.6421372 immediately following publication indefinitely for anyone who wishes to access the data for any purpose.

## Acknowledgements

We thank the Bill & Melinda Gates Foundation for providing funding (INV-001754) to undertake this research.

## Author contributions

Conception: RAC, AP, CKW, RCH, NAM, RGW

Data acquisition and preparation: RAC, AP, CKW, RGW, NAM

Data analysis: RAC, AP, TS, CKW, NAM, RGW

Interpretation of results: RAC, AP, RGW, NAM, TS, KR, SKM, DT, CKW

Manuscript drafting and revisions: RAC, AP, RGW, NAM, TS, KR, SKM, DT, CKW, RB, RCH

All authors had the opportunity to access and verify the data and were responsible for the decision to submit the manuscript for publication.

