## Supplementary Material for "The potential health and economic impacts of new tuberculosis vaccines under varying delivery strategies in Delhi and Gujarat, India: a modelling study"

Rebecca A. Clark et al

**Table of Contents**

SUPPLEMENTARY METHODS 3
1. Summary of tuberculosis in India, Delhi and Gujarat 3
2. Model structure and equations 7
   2.1 Natural history model structure 7
   2.2 Natural history model equations 8
3. Natural history 9
   3.1 Natural history parameter values and data sources 9
   3.2 Operationalising age varying parameters 11
   3.3 Tuberculosis treatment 12
4. Model simulation and calibration 13
   4.1 Model simulation 13
   4.2 Model calibration 13
   4.3 Subnational demography 15
5. Policy scenarios 17
   5.1 No-new-vaccine baseline 17
   5.2 Vaccine delivery 17
      5.2.1 Vaccine scenarios 17
6. Economic analysis methods 19
   6.1 Calculation of disability-adjusted life years 19
   6.2 Tuberculosis-related cost model 19
   6.3 Vaccine introduction costs 19
   6.4 Cost-effectiveness analysis and willingness-to-pay thresholds 21
   6.5 Total costs from the health-system and societal perspectives 22
7. Health impact outcomes 22
SUPPLEMENTARY RESULTS 23
8. No-new-vaccine baseline 23
   8.1 No-new-vaccine baseline calibration 23
   8.2 Posterior distributions 25
9. Health impact results 27
10. Economic results 30
   10.1 Delhi Economic Results - M72/AS01E 30

10.2 Delhi Economic Results - BCG-revaccination 33
10.3 Gujarat Economic Results - M72/AS01$_E$ 36
10.4 Gujarat Economic Results - BCG-revaccination 39
References 42

#### SUPPLEMENTARY METHODS

##### 1. Summary of tuberculosis in India, Delhi, and Gujarat

India is classified as one of the WHO top 30 high tuberculosis burden countries for 2021–2025, in addition to appearing on the high tuberculosis/HIV and drug-resistant tuberculosis lists.[1] The incidence rate of new tuberculosis cases in India in 2021 was estimated at 210 per 100,000 population per year.[2] India is divided into 28 states and 11 union territories, with a total population of over 1.3 billion estimated in 2020 (Figure S1.1).[3] The state with the largest population size is Uttar Pradesh, accounting for approximately 17% of the total population in 2020.[3,4] The rurality of each state varies across India, with almost 90% of the population in 2011 living in a rural area in Himachal Pradesh and Bihar, compared to less than 3% in Delhi and Chandigarh (Figure S1.2).[5]

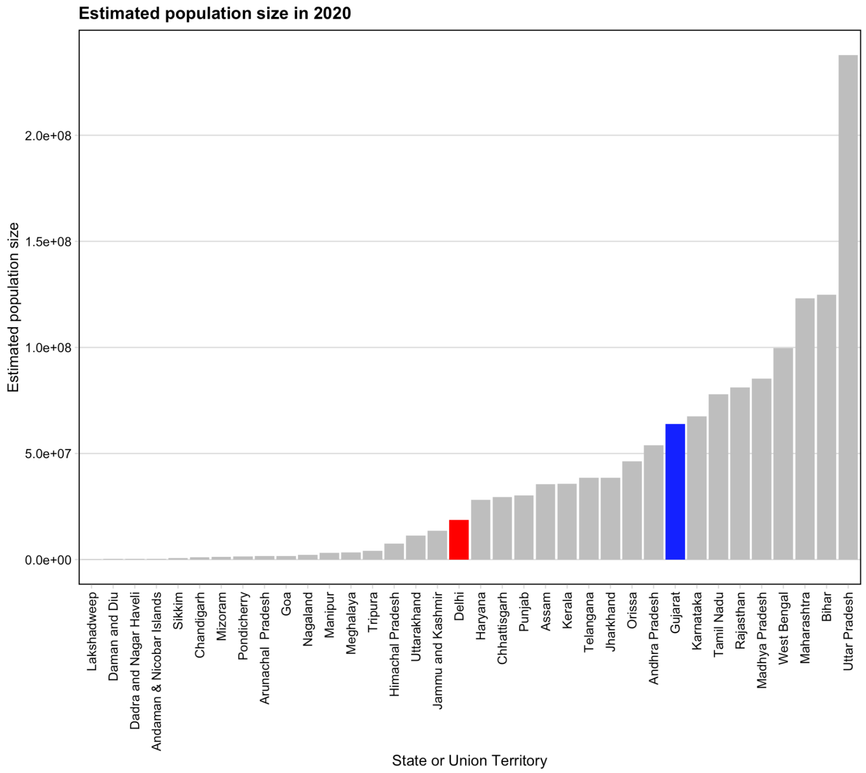


**Figure S1.1** **Estimated population size in 2020 by state and union territory. Delhi is highlighted in red, and Gujarat is highlighted in blue.**[4]

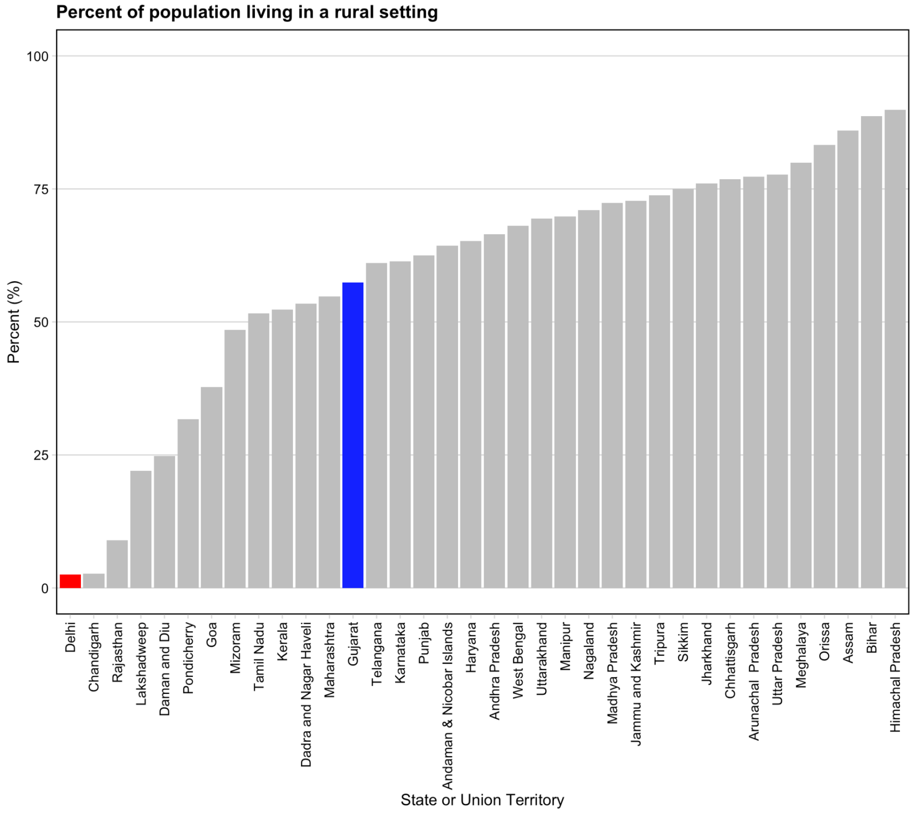


**Figure S1.2 Percent of the population of each state and union territory living in a rural setting in 2011. Delhi is highlighted in red, and Gujarat is highlighted in blue.[5]**

Financial and policy responsibility for the healthcare system falls to the federal government, while the state government is responsible for healthcare delivery.[6] Although tuberculosis treatment is freely available from the public sector, evidence has shown a large proportion of patients are choosing to access care from the private sector.[7–9] A 2019 study from Arinaminpathy et al. estimated that nationally, the percent of treatment months completed in the public sector was 36.0% (33.0, 39.0), ranging from 22.0% (17.0, 25.0) in Bihar to 73.0% (63.0, 79.0) in Himachal Pradesh.[7]

In terms of access to healthcare, there are large variations between and within states depending on the relative proportions of urban and rural communities.[10] States with an increased level of urbanisation have access to options in both the public and private sectors, whereas states are restricted by the limited availability of local healthcare options when rurality is increased.[10] For healthcare services specific to tuberculosis, a systematic review from 2015 investigated the quality of tuberculosis care provided in India, and found that they were often lacking in major areas, including baseline knowledge of tuberculosis symptoms and standard treatment protocol.[11]

The tuberculosis burden varies widely across India (Figure S1.3). The tuberculosis disease prevalence estimate for all ages was estimated at 312 (286–337) per 100,000 in India overall, but ranged by state from 137 (76–198) per 100,000 to 747 (510–984) per 100,000 (almost 5.5 times greater).[12]

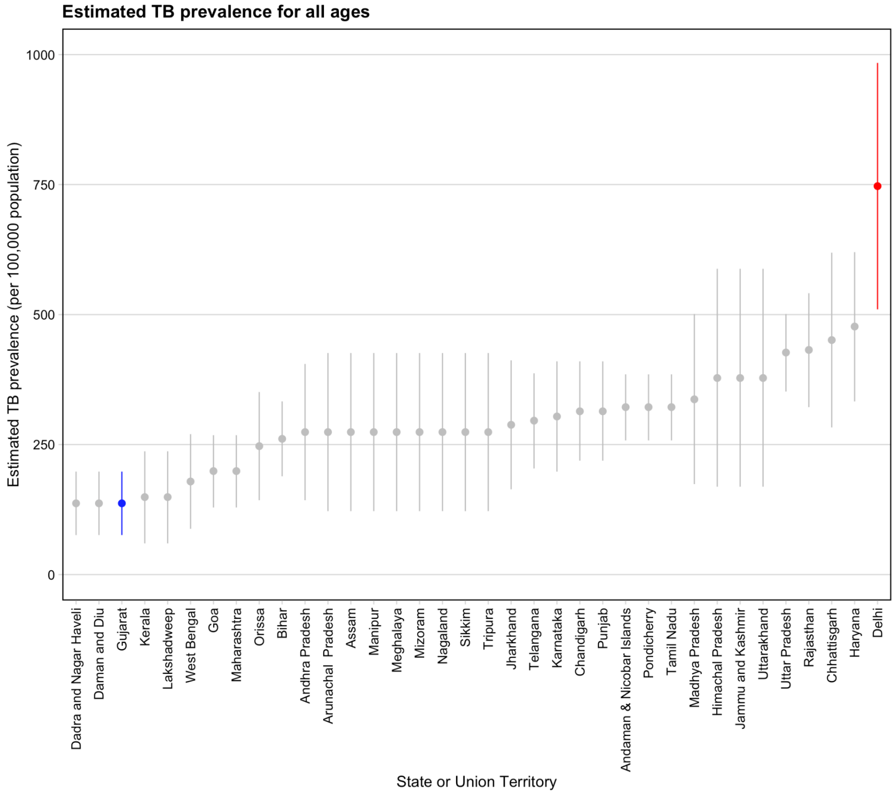


**Figure S1.3 Estimated TB prevalence (per 100,000 population) for all ages by state and union territory.**[12] **Delhi is highlighted in red, and Gujarat is highlighted in blue.**

+ *Gujarat, Dadra and Nagar Haveli, and Daman and Diu were grouped together as one state group for estimating TB prevalence, and therefore have the same estimated value.*

Specifically modelling states and union territories within India will help to support the National Tuberculosis Elimination Program (NTEP) Sub-National Certification of Disease Free Status initiative introduced by the Government of India, which incentivises states and districts to reduce tuberculosis incidence rates. We chose to model Delhi and Gujarat to represent regions with extremes of the wide variation in epidemiology across India (highest and lowest prevalence estimate from the survey respectively), as well as additional distinct characteristics such as population sizes and levels of urbanisation to assess the possible influence of heterogeneity on proposed delivery strategies, as well as to extrapolate to other similar regions.

##### *Delhi*

The National Capital Territory of Delhi, or "Delhi", is a geographically small city and union territory located in the north of India. According to the 2011 census, Delhi had a population of almost 17 million—the 19th largest state or union territory in the country—with 97.5% of the population living in an urban setting.[5] By 2020, the estimated population increased by 12% to almost 19 million.[4] In the recent National TB Prevalence survey in India conducted from 2019-2021, Delhi was estimated to have the highest prevalence per 100,000 population for adults at 534 (365–704) per 100,000 and the highest estimated tuberculosis prevalence for all ages at 747 (510–984) per 100,000.[12] In 2020, Delhi reported over 100,000 tuberculosis cases, with 35% of reported notifications seeking care in the private sector.

***Gujarat***

The state of Gujarat is located on the west coast of India. With an estimated 64 million people living in Gujarat in 2020, it is the 9th largest state by population.[4] Gujarat is increasingly becoming more urban, with around 50% of the population living in urban settings. In contrast to Delhi, Gujarat has one of the lowest estimated tuberculosis prevalence per 100,000 population for adults [141 (78–203) per 100,000], the lowest estimated tuberculosis prevalence per 100,000 population among all ages [137 (76–198) per 100,000], as well as the lowest estimate of the prevalence to notification ratio (0.91).[12] In 2020, Gujarat reported almost 145,000 tuberculosis cases, with 65% of reported notifications seeking care in the public sector. Gujarat was awarded Bronze in 2021 for reducing the incidence rate by 20% compared to 2015 estimates, and it has been reported that six districts out of 26 total have already made claims of disease free status under the Certification of Disease Free Status initiative.

#### 2. Model structure and equations

We extended existing age-stratified compartmental differential equation models of tuberculosis, including dimensions for age, tuberculosis natural history and vaccination.[13,14] As in Clark et al., 2023, we modelled single age groups from age 0 through 79, one compartment for age 80–89 and one compartment for age 90–99. The natural history model structure (Section 2.1) and natural history model equations (Section 2.2) are identical to the supplementary material in Clark et al, 2023 and reproduced here without modification.

##### 2.1 Natural history model structure

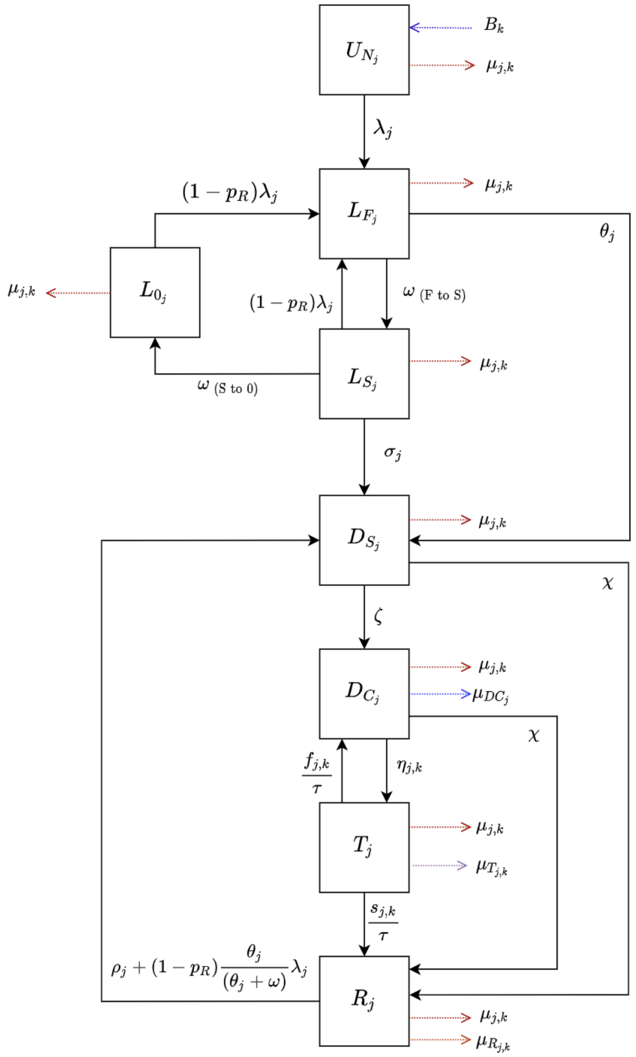


**Figure S2.1 Tuberculosis natural history model structure**

*Subscript j represents parameters that vary by age, and subscript k represents parameters that vary over time.*

*Abbreviations: $U_N$ = Uninfected-Naive; $L_F$ = Latent-Fast; $L_S$ = Latent-Slow; $L_0$ = Latent-Zero, $D_S$ = Subclinical Disease; $D_C$ = Clinical Disease; T = On-Treatment; R = Recovered.*

A natural history structure with eight compartments in Figure S2.1 was created by adapting features of previous models, and has been described previously. The latency structure in this model demonstrates a progressive loss of ability to reactivate, with the reactivation rate in the Latent-Fast compartment greater than in Latent-Slow and greater still than in Latent-Zero, where we assume the rate of reactivation is 0. We do not explicitly have a self-clearance compartment. We assume that those in Latent-Fast can only fast progress to subclinical disease, or continue to remain latent and transition to Latent-Slow. There is no direct transition from Latent-Fast to Latent-Zero.

#### 2.2 Natural history model equations

$Age\ j = 0$

$$\frac{dU_{N_j}}{dt} = B_k - (\lambda_j + \mu_{j,k})U_{N_j}$$

$Age\ j \neq 0$

$$\frac{dU_{N_j}}{dt} = -(\lambda_j + \mu_{j,k})U_{N_j}$$

$$\frac{dL_{F_j}}{dt} = \lambda_j U_{N_j} + (1 - p_R)\lambda_j L_{0_j} + [(1 - p_R)\lambda_j]L_{S_j} - (\omega_{FS} + \theta_j + \mu_{j,k})L_{F_j}$$

$$\frac{dL_{S_j}}{dt} = \omega_{FS} L_{F_j} - (\omega_{S0} + \sigma_j + (1 - p_R)\lambda_j + \mu_{j,k})L_{S_j}$$

$$\frac{dL_{0_j}}{dt} = \omega_{S0} L_{S_j} - [(1 - p_R)\lambda_j + \mu_{j,k}]L_{0_j}$$

$$\frac{dD_{S_j}}{dt} = \theta_j L_{F_j} + \sigma_j L_{S_j} + [\rho_j + (1 - p_R)\frac{\theta_j}{\theta_j + \omega}\lambda_j]R_j - (\chi + \zeta + \mu_{j,k})D_{S_j}$$

$$\frac{dD_{C_j}}{dt} = \zeta D_{S_j} + \frac{f_{j,k}}{\tau}T_j - (\chi + \eta_{j,k} + \mu_{DC_j} + \mu_{j,k})D_{C_j}$$

$$\frac{dT_j}{dt} = \eta_{j,k} D_{C_j} - \left(\frac{s_{j,k} + f_{j,k}}{\tau} + \mu_{T_{j,k}} + \mu_{j,k}\right)T_j$$

$$\frac{dR_j}{dt} = \frac{s_{j,k}}{\tau}T_j + (D_{S_j} + D_{C_j})\chi - [\rho_j + (1 - p_R)\frac{\theta_j}{\theta_j + \omega}\lambda_j + \mu_{R_j} + \mu_{j,k}]R_j$$

#### 3. Natural history

##### 3.1 Natural history parameter values and data sources

Parameters used in the natural history model structure are provided in Table S3.1 below, along with their definitions, sources, and information on whether the parameter is fixed or varied (as well as whether they are varied by age or time) during calibration. Further details about how the age varying parameters are implemented are provided in section 3.2, and further details on parameters related to treatment are provided in section 3.3. The parameter ranges provided for the tuberculosis natural history parameters are priors fitted during calibration in a Bayesian analysis. We assume that all values within the prior range are equally likely. For certain natural history parameters that we believe will not vary within the country, we used the posterior distributions (95% uncertainty intervals) from the National India modelling study from Clark et al., 2023 as prior distributions for subnational modelling. The prior distributions for the National India model from Clark et al, 2023 were pre-specified based on literature review and reviewed as new data became available. Unless otherwise specified, we assume the same ranges for both Delhi and Gujarat.

**Table S3.1 Description of natural history parameters used during calibration for Delhi and Gujarat**

| Description | Units | Symbol | Prior | Fixed or Varying During Calibration | Age Varying | Time Varying | Source |
|---|---|---|---|---|---|---|---|
| ***Births and deaths (excluding on-treatment mortality)*** | | | | | | | |
| Birth rate | Per year | $B_k$ | Population estimates and projections as described in Section 4.3 | Fixed | No | Yes | 5,15 |
| Background mortality rate | Per year | $\mu_{j,k}$ | Calculated in the model from population estimates and projections | Fixed | Yes, age specific mortality rates from demographic dataset | Yes | 5,15 |
| Mortality rate for clinical tuberculosis disease | Per person per year | $\mu_{DC_j}$ | (0.124–0.177) | Varying | Yes, value for children is greater than value for adults | No | Posterior from the National India model[13] |
| Mortality rate post-tuberculosis disease | Per person per year | $\mu_{R_j}$ | 0.22 × [(0.004–0.02)] | Fixed relationship | Yes because $\mu_{j,k}$ varies | Yes because $\mu_{j,k}$ varies | Posterior from the National India model[13] |
| ***Natural History*** | | | | | | | |

| | | | | | | | |
|---|---|---|---|---|---|---|---|
| Force of infection | Per year | $\lambda_j$ | Fitted | Fixed Equation | Yes, age specific contact rates[16] | No | Calculated |
| Probability of transmission per infectious contact | - | $p_T$ | (0–0.0068) | Varying | No | No | Assumed |
| Fraction of total tuberculosis that is extrapulmonary | - | $ep$ | Delhi: 0.440<br>Gujarat: 0.179 | Fixed | No | No | 17–23 |
| Infectiousness of subclinical relative to clinical tuberculosis | - | $r$ | 0.83 | Fixed | No | No | 24 |
| Rate of fast progression to disease, by age | Per person per year | $\theta_j$ | (0.092–0.110) | Varying | Yes; Retain if value for children is less than value for adults. | No | Posterior from the National India model[13] |
| Rate from $L_F$ to $L_S$ | Per person per year | $\omega_{FS}$ | 0.5 | Fixed | No | No | Defined |
| Rate of reactivation from $L_S$, by age | Per person per year | $\sigma$ | (0.00069–0.00112) | Varying | Yes; Retain if value for children is less than value for adults. | No | Posterior from the National India model[13] |
| Rate from $L_S$ to $L_0$ | Per person per year | $\omega_{S0}$ | (0.026–0.037) | Varying | No | No | Posterior from the National India model[13] |
| Rate of progression from $D_S$ to $D_C$ | Per person per year | $\zeta$ | (0.758–1.331) | Varying | No | No | Posterior from the National India model[13] |
| Rate of natural cure from $D_C$ and $D_S$ | Per person per year | $\chi$ | (0.109–0.188) | Varying | No | No | Posterior from the National India model[13] |
| Rate of relapse from R, by age | Per person per year | $\rho_j$ | (0.015–0.023) | Varying | Yes; Retain if value for children is less than value for adults. | No | Posterior from the National India model[13] |
| ***Protection Parameters*** | | | | | | | |
| Protection from reinfection $L_S$, $L_F$, $L_0$, R | - | $p_R$ | (0.616–0.779) | Varying | No | No | Posterior from the National India model[13] |

##### 3.2 Operationalising age varying parameters

We assume that aspects of tuberculosis natural history and mortality vary by age as in Clark et al., 2023.[13] This is implemented by stratifying certain natural history parameters by age and applying age-specific prior ranges and relative constraints during calibration.[25] The following table describes the method used to operationalise the age varying differences in parameters between adults, defined as all ages greater than and equal to 15, and children, defined as all ages less than 15. For the rate per year of reactivation, relapse, and fast progression to tuberculosis disease, we assume that the rate for children is less than that for adults. For mortality rates, we assume the opposite: the rate for children is higher than that for adults.

**Table S3.2 How age varying parameters are operationalised**

| Parameter | Range | Age Varying Description | Age Scaling Parameter | Adults ($\theta_{A15}$) | Children ($\theta_{A0}$) |
|---|---|---|---|---|---|
| $\theta_j$ Rate per year of fast progression | (0.092–0.110) | Retain if value for children is **less** than value for adults | Sample $j_1$ from (0.073–0.66) | Sample $\theta_{A15}$ from (0.092–0.110) | $\max(0.092, \theta_{A15} \times j_1)$ |
| $\sigma_j$ Rate per year of reactivation | (0.00069–0.00112) | Retain if value for children is **less** than value for adults | Sample $j_2$ from (0.340–0.962) | Sample $\sigma_{A15}$ from (0.00069–0.00112) | $\max(0.00069, \sigma_{A15} \times j_2)$ |
| $\rho_j$ Rate per year of relapse | (0.015–0.023) | Retain if value for children is **less** than value for adults | Sample $j_3$ from (0.371–0.969) | Sample $\rho_{A15}$ from (0.015–0.023) | $\max(0.015, \rho_{A15} \times j_3)$ |
| $\mu_{DC_j}$ Clinical TB mortality rate per year | (0.124–0.177) | Retain if value for children is **greater** than value for adults | Sample $S_{Age}$ from (0.597–0.967) | $\mu_{DC_{A0}} \times S_{Age}$ | Sample $\mu_{DC_{A0}}$ from (0.124–0.177) |
| $\mu_{T_j} = \frac{\kappa_j}{\tau}$ On-treatment mortality rate per year | DEL: (0–0.244) GUJ: (0–0.283) | Retain if value for children is **greater** than value for adults | Sample $S_{Age}$ from (0.597–0.967) | $\frac{\kappa_{A0}}{\tau} \times S_{Age}$ | Sample $\kappa_{A0}$ from: Delhi: (0–0.122) Gujarat: (0–0.142) |

##### 3.3 Tuberculosis treatment

Steps for calculating treatment initiation, treatment completion, non-completion, and mortality rates are described in the Supplementary Material for Clark et. al, 2023, with Delhi and Gujarat specific adjustments described below.

***Treatment initiation***

We assumed that the steps for treatment initiation in Delhi were identical to Clark et al., 2023. We allowed the upper bound of the prior range for the treatment initiation rate in 2019 (eta) for Gujarat to be extended from 1 to 2 to allow for greater healthcare seeking (more than 100% of those with prevalent tuberculosis to be treated within one year).

***Treatment outcomes***

Region-specific treatment completion, non-completion, and mortality fractions were calculated as a weighted average of public and private sector reported estimates from the India TB Reports from 2018–2021.[21–23,26] We used the India TB Report 2022 to determine how many notifications were expected to be reported from the public and private sector for Delhi and Gujarat, and determined the proportion of treatment expected to occur in each sector (Delhi: 73% in the public sector, 27% in the private sector; Gujarat: 66% in the public sector, 34% in the private sector). We assumed that this proportion was constant over time. We then calculated the fraction of treatment completion, non-completion, and mortality for each region for the public and private sector separately, and then as a weighted average to obtain one estimate of each outcome for each region.

The weighted average of the treatment completion and non-completion estimates were used to calculate the SFR, which represented the ratio between treatment completions to the sum of treatment completions and non-completions. This was estimated to be 0.941 in Delhi and 0.949 in Gujarat. The weighted average of the on-treatment mortality was multiplied by 2 to give an upper bound of the range for kappa. This was estimated to be 0.122 in Delhi and 0.142 in Gujarat.

**Table S3.3 Calculating treatment outcome parameter values for adults and children**

| Parameter | Adults | Children |
|---|---|---|
| $\kappa_j$ On-treatment mortality fraction | $\kappa_{A0} \times S_{Age}$ | Sample $\kappa_{A0}$ from:<br>Delhi: (0–0.122)<br>Gujarat: (0–0.142) |
| $s_j$ On-treatment completion fraction | $(1-\kappa_{A15})\mathrm{SFR}$ | $(1-\kappa_{A0})\mathrm{SFR}$ |
| $f_j$ On-treatment non-completion fraction | $(1-\kappa_{A15})(1-\mathrm{SFR})$ | $(1-\kappa_{A0})(1-\mathrm{SFR})$ |

### 4. Model simulation and calibration

#### 4.1 Model simulation

Model simulation was as in both Clark et al., 2023 studies, reproduced here with some small modifications.[13,14] We specified a system of ordinary differential equations defining the derivatives with respect to time of a set of state variables, to simulate the tuberculosis epidemic between 1900 and 2050. We initialised the simulation by distributing the population between the eight tuberculosis natural history states using a fitted parameter representing the proportion of the population uninfected at the start of the simulation. For each year of the simulation (1900–2050), our models are designed to exactly match the age-specific population estimates and projections.

#### 4.2 Model calibration

For this subnational modelling analysis of Delhi and Gujarat, we followed the same modelling approach as in both Clark et al., 2023 studies, reproduced here with some small modifications.[13,14]

Broadly, this was as follows:

1. Construct a mechanistic model
2. Calibrate the model by identifying areas of the input parameter space where the output of the mechanistic model was consistent with the historical epidemiologic data
3. Use the calibrated model to simulate and predict future tuberculosis epidemiology and model new vaccines

In the context of this analysis, step 1 was achieved by creating the compartment differential equation model as specified in Section 2. For step 2, we independently calibrated a model by identifying areas of the parameter space that made the output of the model match the corresponding calibration targets (Table S4.1). Further details on the sources for the calibration targets and any additional modifications are in the subsequent sections.

The model was fitted to the calibration targets using history matching with emulation, a method that allows us to explore high-dimensional parameter spaces efficiently and robustly.[27–30] History matching progresses as a series of iterations, called waves, where implausible areas of the parameter space, i.e., areas that are unable to give a match between the model output (e.g., the predicted disease prevalence by the model) and the empirical data (e.g., the disease prevalence calibration target from the National TB Prevalence Survey), are found and discarded. In order to identify implausible parameter sets, emulators, which are statistical approximations of model outputs that are built using a modest number of model runs, are used. Emulators provide an estimate of the value of the model at any parameter set of interest, with the advantage that they are orders of magnitude faster than the model.

History matching with emulation, implemented through the *hmer* package in R,[31,32] considerably reduced the size of the parameter space to investigate. Rejection sampling was then performed on the reduced space to identify at least

1000 parameter sets that matched all targets. Once we had obtained 1000 parameter sets that produced output consistent with the calibration targets, we used those parameter sets with the mechanistic model to simulate the future (step 3) for each region.

We calibrated Delhi to three calibration targets and, separately, Gujarat to four calibration targets based on the differences in regionally available data. Calibration targets are in Table S4.1 below.

**Table S4.1 Calibration targets for Delhi and Gujarat**

| Calibration Target | Year | Delhi | Gujarat |
|---|---|---|---|
| Tuberculosis disease prevalence for all ages (per 100,000 population)[12] | 2021 | 747 (510–984) | 137 (76–198) |
| Tuberculosis disease prevalence for adults (per 100,000 population) | 2011 | *NA* | 383 (315–451) |
| Tuberculosis case notification rate for all forms and all ages (per 100,000 population) with 20% bounds[33] | 2021 | 536 (429–644) | 137 (110–165) |
| Subclinical TB prevalence ratio[34] | 2020 | 0.564 (0.428–0.685) | |

***Adjustments to calibration targets***

The notification rate from the India TB Report 2022 for Gujarat was 204 per 100,000 population. When comparing this estimate to the disease prevalence estimate from the National Tuberculosis Prevalence survey, a higher rate of the population was treated for tuberculosis than currently had prevalent disease (204 per 100,000 notifications compared to 137 per 100,000 with prevalent disease). We know that when healthcare services are improved and the prevalence of tuberculosis decreased, more false positives are expected. Therefore, we adjusted the notification rate target in Gujarat down to account for the possibility of false positives. As only 35% of the reported notifications in Gujarat were bacteriologically confirmed, we adjusted the reported notification rate (204 per 100,000 population) relative to the proportion of reported notifications that were bacteriologically confirmed in Delhi (52%), to obtain a new case notification target of 137 per 100,000.

As described in Section 3, we also allowed healthcare seeking to increase in Gujarat, by increasing the "eta" parameter to allow more than 100% of those with prevalent disease to be treated within one year. We included both adjustments (adjusting the case notification rate target down and increasing the treatment seeking parameter) as it is unknown which is correct, and we allowed the model to determine the best fit. We do not believe these modifications would be representative of Delhi, and therefore are only included for Gujarat.

#### 4.3 Subnational demography

United Nations Population Data and Projections were available for India overall for single ages and years from 1950–2100, but this level of detailed data was not available for Delhi and Gujarat. We combined all available data to ensure we represented the total population size and age distribution as accurately as possible, as these two aspects may play an important role in vaccine impact estimation.

##### *Total Population Size*

To obtain accurate representations of total population size, we first collated all available demographic data for Delhi, and Gujarat. From the Government of India Census data, we obtained single age numbers (1000s) in 1991, 2001 and 2011. From the most recent Government of India census (2011), we obtained single age projections (1000s) for ages 5 to 23 in years 2016, 2021, 2026, 2031, and 2036, 5 year age group projections (1000s) in years 2016, 2021, 2026, 2031, and 2036 and total population projections (1000s) in years 2011 to 2036.

The total population estimates and projections for Delhi and Gujarat used in the model simulation are shown in Figure S4.1. Total population estimates were available from census data in 1991, 2001, 2011, and total population projections were available for 2011–2036. We used a linear interpolation between the estimates in 1991 and 2001 for the years in between, and similarly, a linear interpolation between the estimates in 2001 and 2011 for the years in between. These data and projections are represented on Figure S4.1 with the red and blue lines for Delhi and Gujarat respectively. The dashed grey lines represent projecting backwards and forwards from the data by holding the ratio between the population in Delhi or Gujarat and the population in India constant. To explain the method in further detail, the earliest data point available was the total population size in 1991. Dividing the total population size in Delhi or Gujarat by the total population size in India overall gives us a ratio we call $P_D$ and $P_G$ respectively. We then multiplied the population size in India from 1950–1990 by these ratios to obtain an estimate of the total population size in Delhi and Gujarat. We used the same method with the latest available projection (2036) to project forward.

##### *Age Distribution*

To accurately represent the age distribution in Delhi and Gujarat, we compared the age distribution projections in 2011, 2026, and 2036 for India, Delhi, and Gujarat from the 2011 census (Figure S4.2). We assumed that the distribution was similar enough to use the same age composition for Gujarat as in India, but observed a higher proportion of adults in Delhi. Therefore, the age distribution in 2011 for Delhi was applied to the total estimated population for all years leading up to 2011, and similarly, the age distribution in 2036 was applied for all years projecting forward from 2036. For the years between 2011 and 2036, we applied a linear interpolation between the age compositions in 2011 and 2026, and the age compositions in 2026 and 2036.

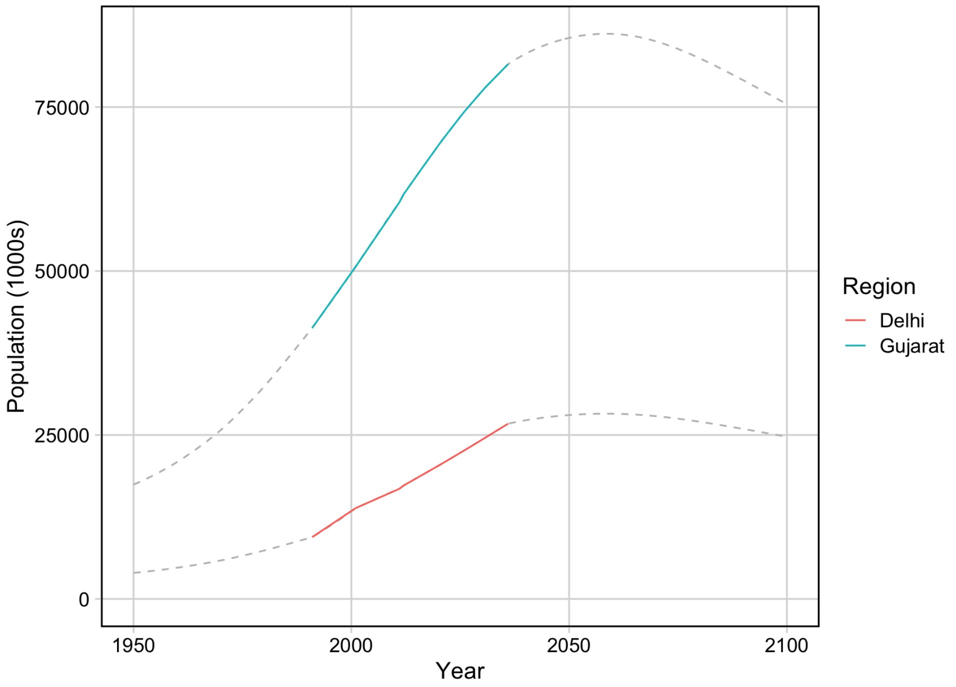


**Figure S4.1** **Total population estimates and projections for Delhi and Gujarat used in model simulation**

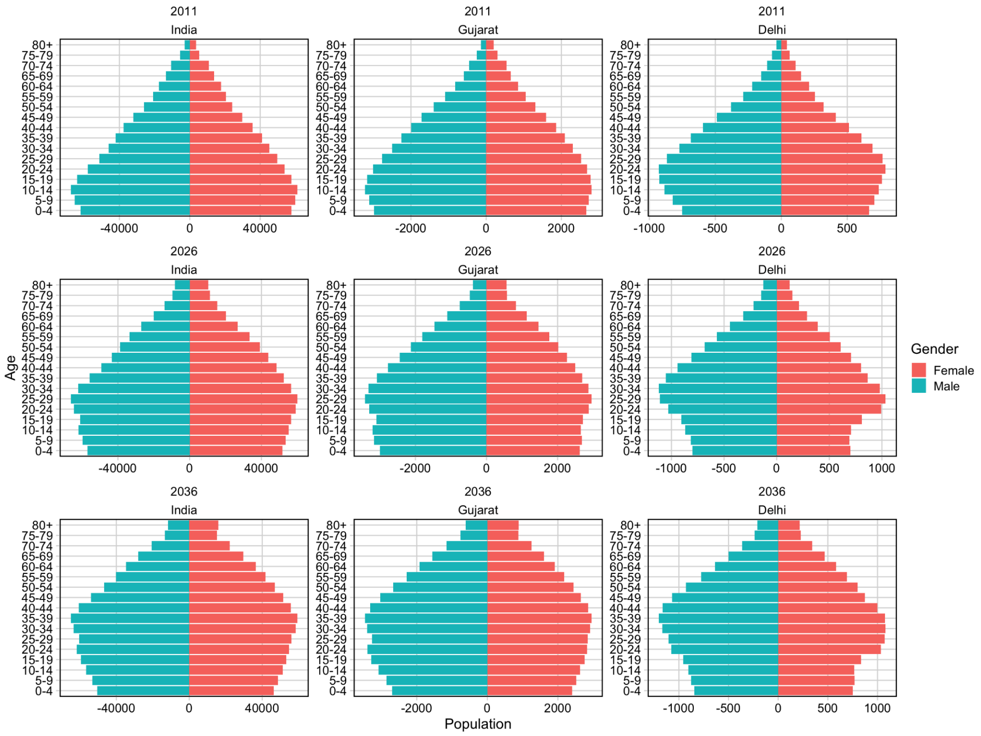


**Figure S4.2** **Age structure in 2011, 2026, and 2036 for India, Gujarat, and Delhi**

#### 5. Policy scenarios

Methods for introducing policy scenarios in Delhi and Gujarat are as in Clark et al., 2023, reproduced here with some small modifications.[13]

##### 5.1 No-new-vaccine baseline

The primary no-new-vaccine simulated was the no-new-vaccine baseline, which assumed non-vaccine tuberculosis interventions continue at current levels into the future. As reported country-level data includes the high coverage levels of neonatal BCG vaccination, this was not explicitly modelled. We assumed that BCG vaccination would not be discontinued over the model time horizon.

##### 5.2 Vaccine delivery

Two recently completed phase IIb trials have demonstrated encouraging efficacy results. The $M72/AS01_E$ candidate vaccine is a subunit vaccine for which results from a completed Phase IIb trial were published at the end of 2019.[35] After three years of follow-up, the efficacy of $M72/AS01_E$ at preventing disease in latently infected adults from South Africa, Zambia, and Kenya was estimated at 49.7% (95% confidence interval = 2.1–74.2).[35] To confirm this finding, a larger, Phase III follow-up study which includes participants who are uninfected, adolescents, as well as those living with HIV to assess safety and immunogenicity in these populations, is anticipated to begin in early 2024.

BCG-revaccination (administering a second dose of BCG to those who were vaccinated neonatally) was previously implemented in many countries, however evidence did not support the effectiveness of this practice. Interest in BCG-revaccination has recently been renewed following results from a trial for the vaccine candidate, H4:IC31. BCG-revaccination was assessed as a third parallel arm alongside H4:IC31 and a placebo in a pre-infection population in South Africa, and although neither vaccine appeared efficacious at preventing infection, BCG-revaccination appeared efficacious at preventing sustained infection (defined as three consecutive positive tests after day 84 of the trial) with an efficacy of 45.4% (6.4–68.1).[36] A larger trial of BCG-revaccination versus placebo in 1800 healthy adolescents from across South Africa is now underway to verify this finding.

We evaluated introducing vaccines with $M72/AS01_E$ and BCG-revaccination characteristics compared to the no-new-vaccine baseline as described in the subsequent sections.

###### 5.2.1 Vaccine scenarios

For each vaccine product, we established one *Basecase* vaccine scenario based on clinical trial data and expert opinion. We then varied vaccine product and delivery scenarios as univariate scenario analyses from the *Basecase* scenario as described in Table S5.1. Vaccine delivery assumptions and model structure are identical to those described in Section 4.2 and 4.3 Clark et al., 2023.[13]

**Table S5.1 M72/AS01$_E$ and BCG-revaccination *Policy Scenarios* and *Vaccine Characteristic and Coverage Scenarios* for Delhi and Gujarat**

<table>
<tr><th rowspan="2">Characteristic</th><th colspan="2">M72/AS01<sub>E</sub></th><th colspan="2">BCG-revaccination</th></tr>
<tr><th>Basecase</th><th>Univariate scenario analyses</th><th>Basecase</th><th>Univariate scenario analyses</th></tr>
<tr><td colspan="5">Policy Scenarios</td></tr>
<tr><td>Age targeting</td><td>Routine age 15, campaign for ages 16–34</td><td>Older Ages: Routine age 17, campaign for ages 18–55<br><br>All Adults: Routine age 18, campaign for ages 19+</td><td>Routine age 10, campaign for ages 11–18</td><td>Older Ages: Routine age 15, campaign for ages 16–34<br><br>All Adults: Routine age 18, campaign for ages 19+</td></tr>
<tr><td colspan="5">Vaccine Characteristic and Coverage Scenarios</td></tr>
<tr><td>Efficacy</td><td>50%</td><td>60%, 70%</td><td>45%</td><td>70%</td></tr>
<tr><td>Mechanism of effect</td><td>Prevents disease</td><td>Prevents infection and disease</td><td>Prevents infection</td><td>Prevents infection and disease</td></tr>
<tr><td>Infection status at time of vaccination required for efficacy</td><td>Any infection (current / no current infection)</td><td>Current infection only</td><td>No current infection only</td><td>Any infection (current / no current infection)</td></tr>
<tr><td>Duration of protection</td><td>10 years</td><td>5, 15, 20</td><td>10 years</td><td>5, 15, 20</td></tr>
<tr><td>Introduction year</td><td>2030</td><td>2036</td><td>2025</td><td>2031</td></tr>
<tr><td>Achieved coverage</td><td>Medium:<br>80% age 15, 70% campaign</td><td>Low:<br>70% routine,<br>50% campaign<br><br>High:<br>90% routine,<br>90% campaign</td><td>Medium:<br>80% routine, 80% campaign</td><td>Low:<br>70% routine, 70% campaign<br><br>High:<br>90% routine, 90% campaign</td></tr>
</table>

#### 6. Economic analysis methods

We used the same economic analysis methods as in Clark et al., 2023, reproduced here with minor modifications.[13] Before undertaking this work, we established an economic analysis plan, involving stakeholders and government officials to ensure we had incorporated all necessary information and planned to report on all key outcomes, to outline the methods used in this work. This is summarised below.

##### 6.1 Calculation of disability-adjusted life years

We calculated the difference in total disability-adjusted life years (DALYs) from vaccine introduction to 2050 for each scenario compared to the no-new-vaccine baseline. We used the disability weight for tuberculosis disease from the Global Burden of Disease 2019 study,[37] and age-specific life expectancy estimates for India overall from the United Nations Development Programme.[38] To incorporate parameter uncertainty in years lost due to disability weight estimates, we made 1000 draws from disability weight uncertainty ranges.

##### 6.2 Tuberculosis-related cost model

We estimated health system unit costs, patient costs and productivity losses based on a scoping review of published literature. For the tuberculosis programme, we obtained unit costs for drug-susceptible and drug-resistant tuberculosis treatment and diagnostic costs, which are provided in Table S6.1. Uncertainty in cost estimates is characterised through gamma distributions around plausible unit cost estimates in a probabilistic sensitivity analysis.

##### 6.3 Vaccine introduction costs

There was considerable uncertainty in the cost of delivering a vaccine, including the price of vaccine compounds and programmatic delivery among adolescents. Based on expert opinion from funders, for the M72/AS01E vaccine we assume a $2.50 per-dose vaccination price with two doses per course assumed in the Basecase. Based on the average estimated BCG price from 2020–2023 from UNICEF,[39] the vaccine price per dose for BCG-revaccination was set at $0.17, with one dose assumed per course.

Due to uncertainty in unit costs of vaccine supply and introduction among populations who may not typically receive large-scale mass vaccination, we make several assumptions around costs to supply and introduction of vaccines. One-time vaccine introduction costs are included in years where there is a campaign and represent non-recurring costs such as establishing infrastructure and providing training for healthcare professionals. The costs were assumed to be $2.40 (1.20–4.80) per total targeted age group population size (as opposed to the actual number of recipients) based on the vaccine introduction support policy of Gavi, the Vaccine Alliance.[40] Vaccine delivery was assumed to be $2.50 (1.00–5.00) per dose, with a further $0.11 (0.06–0.22) supply costs per dose.

In Clark et al., 2023, the cost of recipient vaccination time for India was $0.94 (0.13–1.52), which was calculated by multiplying a wage proxy of GDP per capita for India by an estimate of the time required for vaccination. To represent potential differences in the cost of recipient vaccination time between Delhi and Gujarat due to differences in urban and rural access to healthcare, we included a multiplier on the cost of recipient vaccination time estimate for Delhi which was informed by the average distance to a health facility from the District Level Household and Facility Survey 2007-08.[41] The average distance to a health facility for Gujarat was similar to India overall, and therefore we used the same estimate. For Delhi, the average distance to a health facility was much lower, and therefore we included a multiplier equal to 0.308 on the sampled estimate. We assume a 5% wastage rate.

For each year in the five-year scale up, the vaccination cost is calculated as:
Vaccination cost = (one time introduction costs) × (targeted age group population size)× 0.2 + (number of people vaccinated) × (number of doses) × (vaccine price + vaccine supply costs + cost of delivery) × (1 + wastage)

For each year where there is a repeat campaign, the vaccination cost is calculated as:
Vaccination cost = (one time introduction costs) × (targeted age group population size) + (number of people vaccinated) × (number of doses) × (vaccine price + vaccine supply costs + cost of delivery) × (1 + wastage)

For each year where there is only routine delivery of the vaccine, the vaccination cost is calculated as:
Vaccination cost = (number of people vaccinated) × (number of doses) × (vaccine price + vaccine supply costs + cost of delivery) × (1 + wastage)

For the vaccination cost from the societal perspective, the patient time cost of vaccination is added as a multiplier to the number of doses, and therefore included in the equation along with vaccine price, vaccine supply costs, and the cost of delivery.

##### Table S6.1 Tuberculosis testing, diagnostic, and vaccination related cost inputs

| Unit Cost | Estimate | Lower Bound | Upper Bound | Sources |
|---|---|---|---|---|
| Unit cost of testing/diagnosis for drug-susceptible cases per person | $22.45 | $18.37 | $26.53 | 42 |
| Unit cost of testing/diagnosis for drug-resistant cases per person | $24.36 | $5.04 | $117.81 | 43 |
| Unit cost of treatment for drug-susceptible cases per person | $317.00 | $254.00 | $374.00 | 44 |
| Unit cost of treatment for drug-resistant cases per person | $3,891.00 | $3,382.00 | $4,401.00 | 45 |
| Non-medical patient cost per drug-susceptible tuberculosis disease episode (including transportation) per person | $51.25 | $22.12 | $76.94 | 46,47 |
| Indirect patient cost per drug-susceptible tuberculosis disease episode (time spent on treatment and transport × wage) per person | $117.01 | $24.04 | $460.24 | 47,48 |
| Non-medical patient cost per drug-resistant tuberculosis disease episode (including transportation) per person | $143.49 | $61.95 | $215.42 | 46,47 |
| Indirect patient cost per drug-resistant tuberculosis disease episode (time spent on treatment and transport × wage) per person | $327.63 | $67.30 | $1,288.66 | 47,48 |
| Recurrent vaccine delivery cost per person per dose | $2.50 | $1.00 | $5.00 | 40 |
| One-time vaccine introduction costs per targeted person | $2.40 | $1.20 | $4.80 | 40 |
| Vaccine supply costs per person per dose | $0.11 | $0.06 | $0.22 | 49 |
| Cost of vaccination time per person per dose | $0.94 | $0.13 | $1.52 | 50,51 |

#### 6.4 Cost-effectiveness analysis and willingness-to-pay thresholds

We calculated the incremental cost effectiveness ratio as the ratio between the incremental benefit, in DALYs averted, and the incremental cost, in USD, for each run across vaccination and baseline scenario. Both costs and benefits were discounted to 2025 (when vaccination began) at 3% per year, per guidelines.[52] We measured cost-effectiveness by 2050 against three India specific cost thresholds: 1x gross domestic product (GDP) per-capita (US$1,928), and two country-level opportunity cost thresholds defined by Ochalek et al [the upper (US$443), and lower (US$328) bounds].[53]

#### 6.5 Total costs from the health-system and societal perspectives

The following costs are included in the health-system perspective:

- Vaccine costs: One-time vaccine introduction costs, recurring vaccine delivery costs, vaccine price per dose, and supply costs
- Cost of testing and diagnosis for drug-susceptible and drug-resistant cases
- Cost of treatment for drug-susceptible and drug-resistant cases

In addition to the costs from the health-system perspective, costs from the societal perspective include:

- Vaccine costs: Patient time cost for vaccination
- Non-medical patient costs (including transportation) for drug-susceptible and drug-resistant cases
- Indirect patient costs for drug-susceptible and drug-resistant cases

### 7. Health impact outcomes

The following measures were calculated for each vaccine scenario as the median and 95% uncertainty range:

- Percent incidence rate reduction in 2050 for each vaccine scenario compared to the estimated value in 2050 by *No-New-Vaccine* baseline
- Percent mortality rate reduction in 2050 for each vaccine scenario compared to the estimated value in 2050 by *No-New-Vaccine* baseline
- Cumulative cases averted for each vaccine scenario between 2025 and 2050 compared to the cumulative number of cases estimated by the *No-New-Vaccine* baseline between the corresponding years
- Cumulative deaths averted for each vaccine scenario between 2025 and 2050 compared to the cumulative number of cases estimated by the *No-New-Vaccine* baseline between the corresponding years
- Cumulative treatments averted for each vaccine scenario between 2025 and 2050 compared to the cumulative number of cases estimated by the *No-New-Vaccine* baseline between the corresponding years

#### SUPPLEMENTARY RESULTS

#### 8. No-new-vaccine baseline

##### 8.1 No-new-vaccine baseline calibration

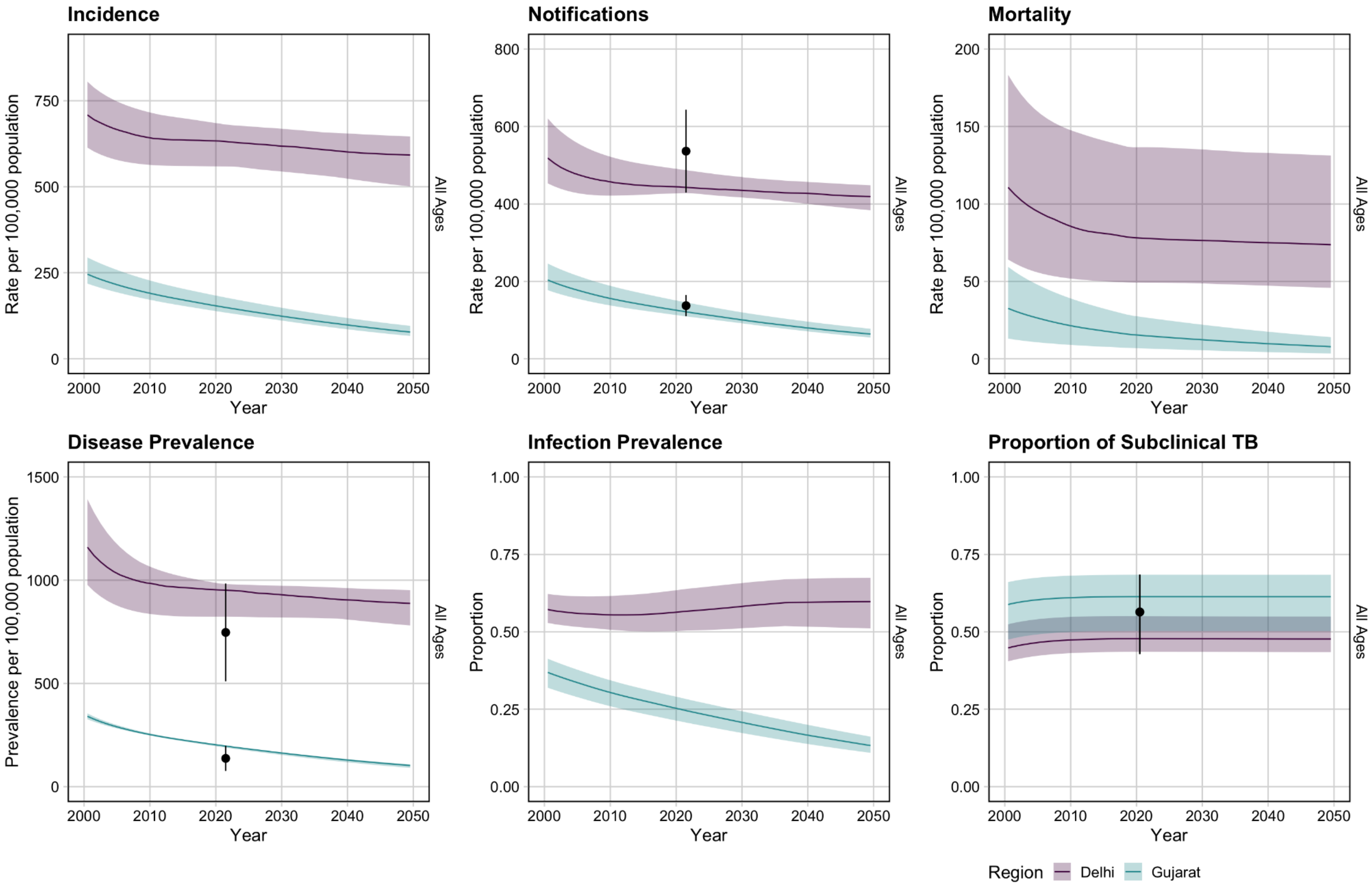


**Figure S8.1 Baseline no-new-vaccine trends from 2000–2050 for all ages for Delhi and Gujarat**

*The trend line indicates the median modelled output with 95% uncertainty in shaded. The black dot and vertical line is the calibration target from Table S4.1.*

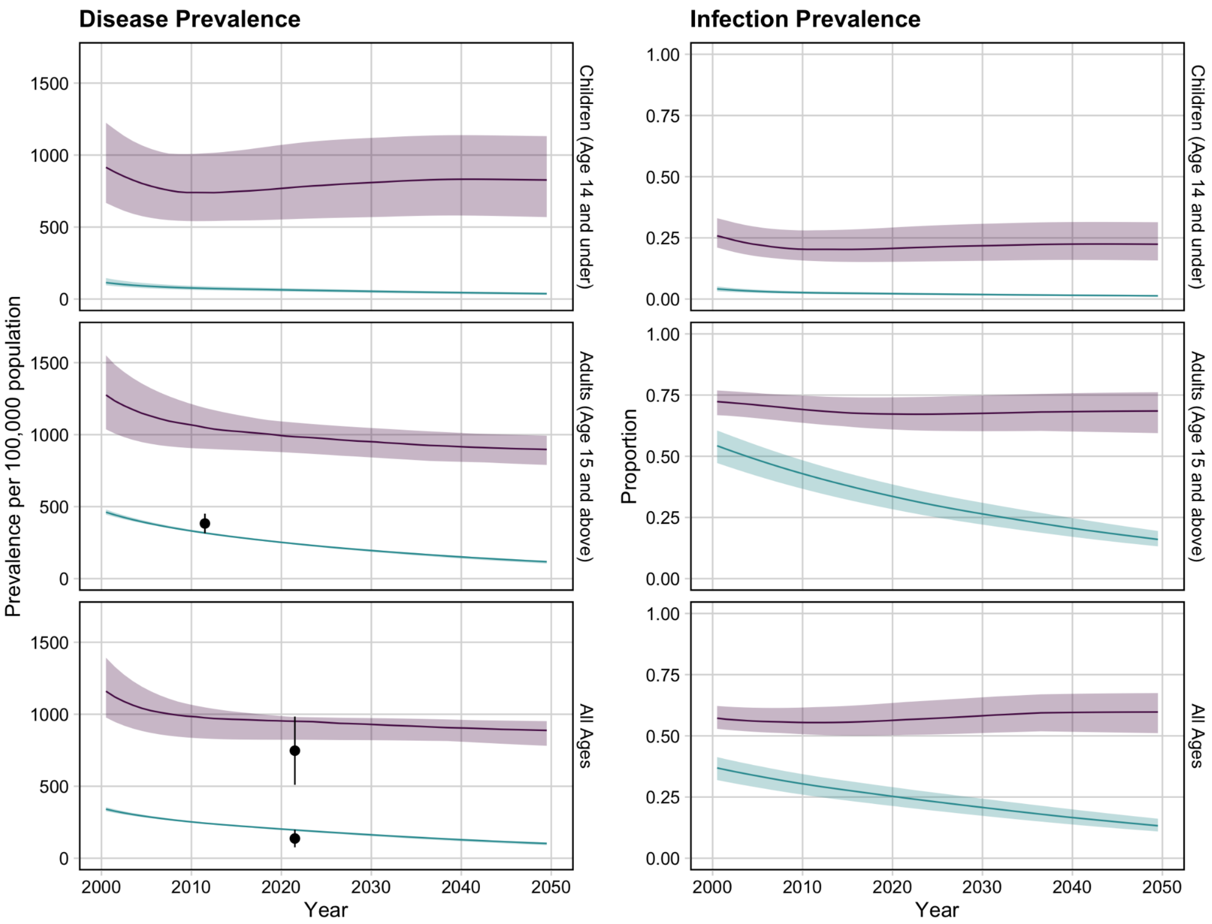


**Figure S8.2 Age-specific trends of tuberculosis disease and infection prevalence in Delhi and Gujarat**

#### 8.2 Posterior distributions

Posterior distribution for the 1000 parameter sets used for vaccine impact estimation are in Figure S8.3 for Delhi and Figure S8.4 for Gujarat.

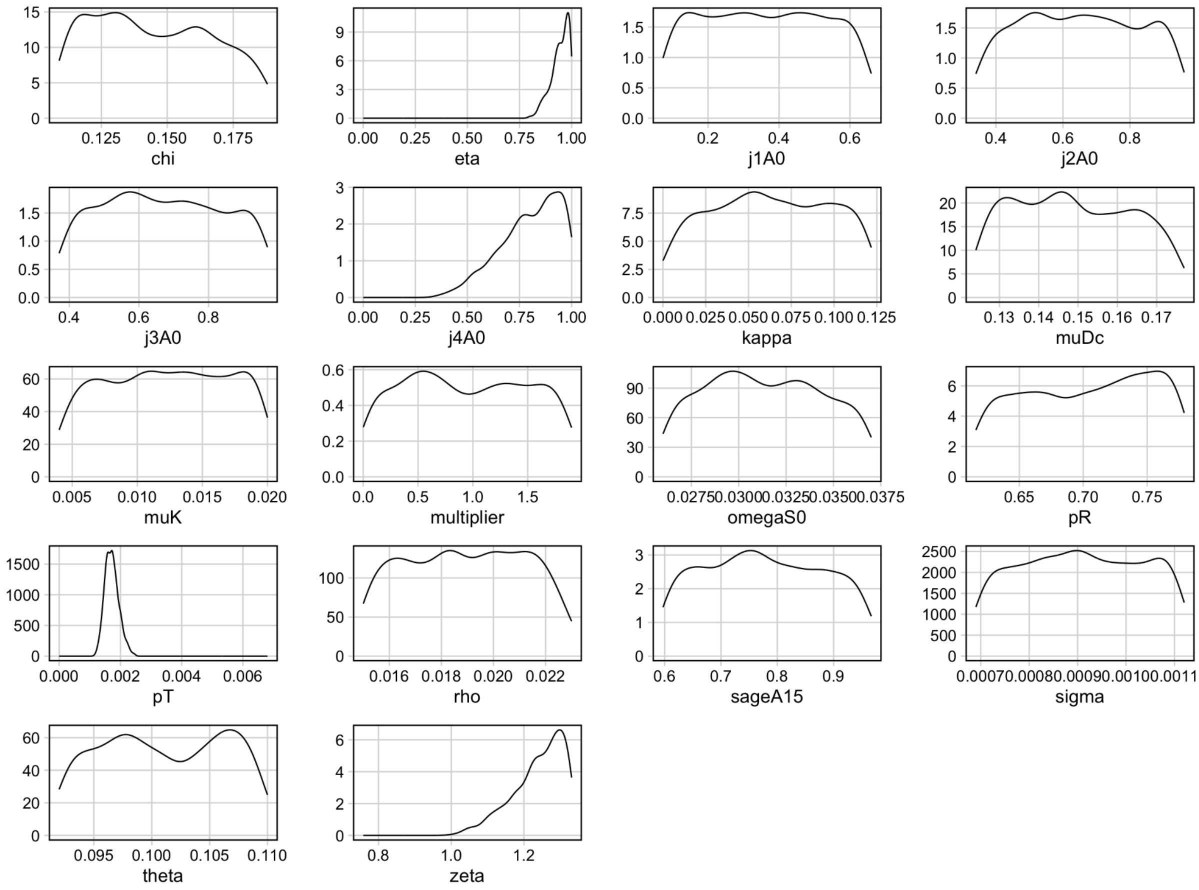


**Figure S8.3 Posterior distributions for the 1000 parameter sets of the 18 parameters varied during calibration for Delhi**

*Parameters are plotted on their prior distributions. Definitions: chi = rate of natural cure, eta = rate of treatment initiation, j1A0 = age multiplier for rate of fast progression (theta), j2A0 = age multiplier for rate of reactivation (sigma), j3A0 = age multiplier for rate of relapse (rho), j4A0 = age multiplier for rate of treatment initiation, kappa = on-treatment mortality fraction, muDc = rate of clinical disease mortality, muK = rate of background mortality for increased mortality rate from the Recovered compartment, multiplier = the multiplier to see the initial distribution of the population into the natural history compartments, omegaS0 = rate of progression between Latent-Slow and Latent-Zero, pR = protection from reinfection for those in the Latency or Recovered compartments, pT = rate of transmission, rho = rate of relapse, sageA15 = age multiplier for mortality rates, sigma = rate of reactivation, theta = rate of fast progression following infection, zeta = rate of progression from subclinical to clinical disease compartments.*

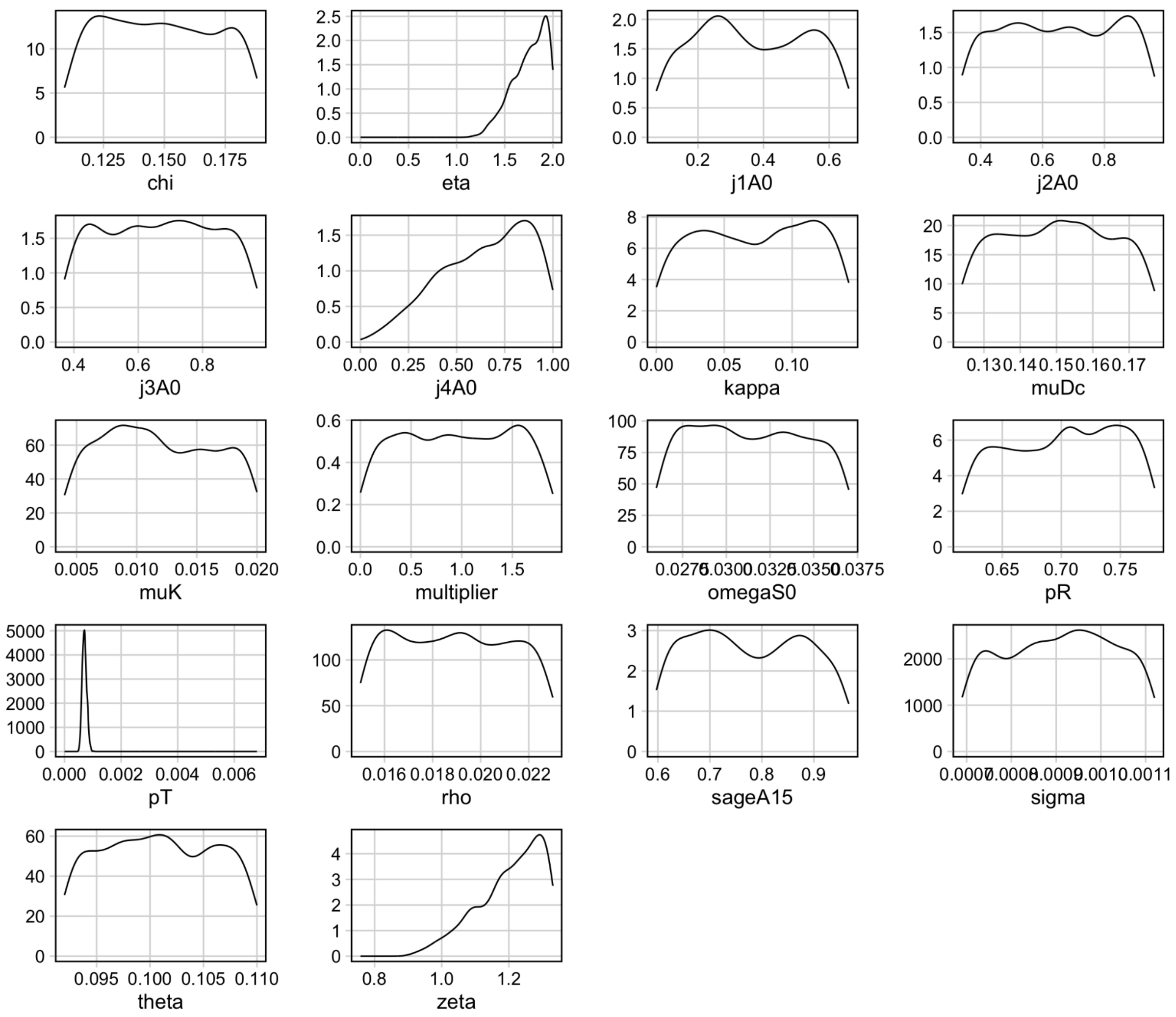


**Figure S8.4 Posterior distributions for the 1000 parameter sets of the 18 parameters varied during calibration for Gujarat**

*Parameters are plotted on their prior distributions. Definitions: chi = rate of natural cure, eta = rate of treatment initiation, j1A0 = age multiplier for rate of fast progression (theta), j2A0 = age multiplier for rate of reactivation (sigma), j3A0 = age multiplier for rate of relapse (rho), j4A0 = age multiplier for rate of treatment initiation, kappa = on-treatment mortality fraction, muDc = rate of clinical disease mortality, muK = rate of background mortality for increased mortality rate from the Recovered compartment, multiplier = the multiplier to see the initial distribution of the population into the natural history compartments, omegaS0 = rate of progression between Latent-Slow and Latent-Zero, pR = protection from reinfection for those in the Latency or Recovered compartments, pT = rate of transmission, rho = rate of relapse, sageA15 = age multiplier for mortality rates, sigma = rate of reactivation, theta = rate of fast progression following infection, zeta = rate of progression from subclinical to clinical disease compartments.*

#### 9. Health impact results

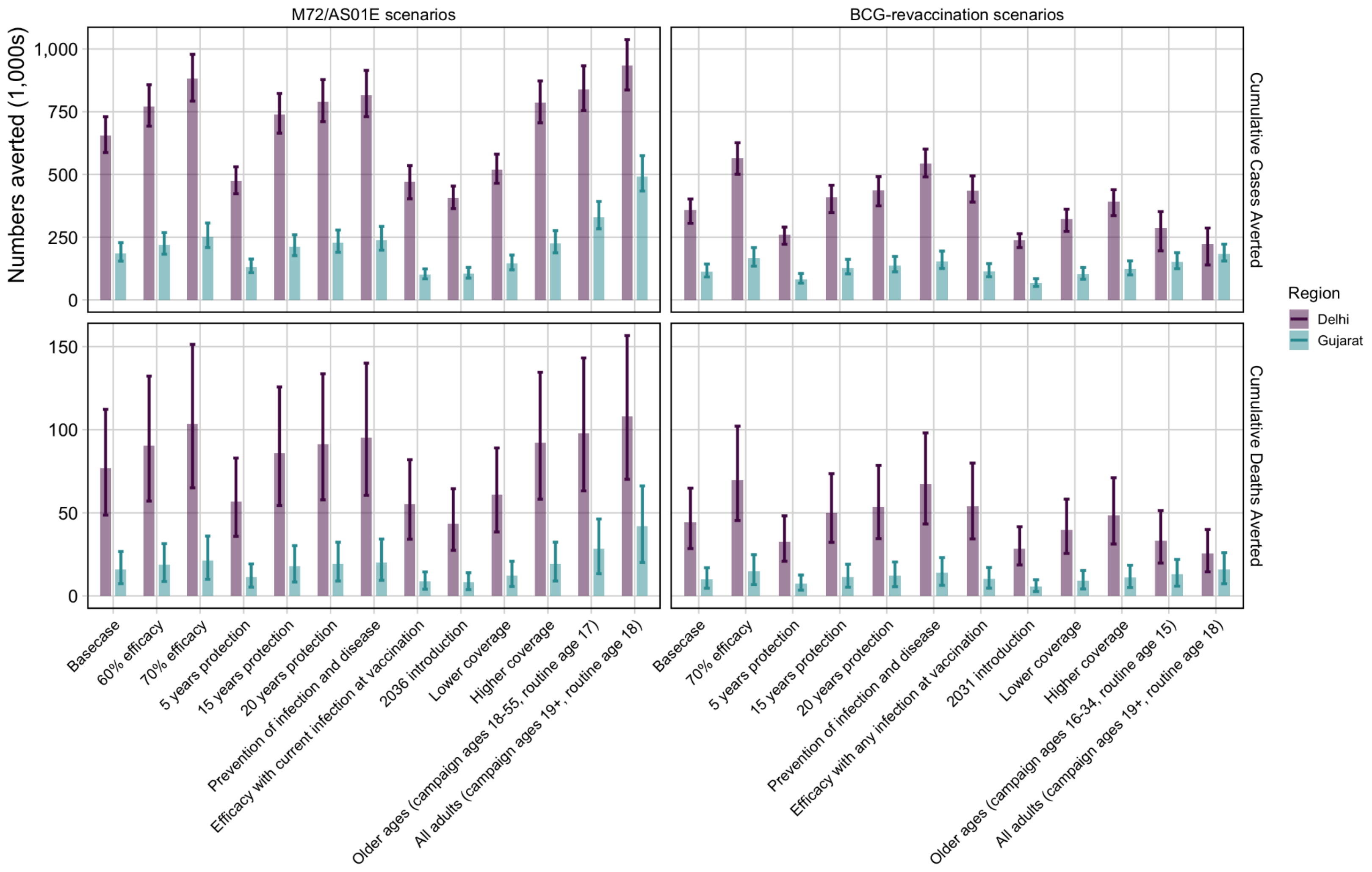


**Figure S9.1 Cumulative cases and deaths averted by vaccine scenarios in Delhi (purple) and Gujarat (blue)**

**Table S9.1** **Cumulative cases and deaths averted between 2025–2050 and rate reductions in 2050 for the vaccine scenarios compared to the no-new-vaccine baseline**

| Scenario | Cumulative cases averted between 2025–2050 (1000s) | | Cumulative deaths averted between 2025–2050 (1000s) | | Incidence rate reduction in 2050 (%) | | Mortality rate reduction in 2050 (%) | |
|---|---|---|---|---|---|---|---|---|
| | Delhi | Gujarat | Delhi | Gujarat | Delhi | Gujarat | Delhi | Gujarat |
| **M72/AS01$_E$ scenarios** | | | | | | | | |
| Basecase | 655.2 (587.4–729.8) | 186.1 (154.6–228.4) | 76.9 (48.7–112.3) | 15.9 (7.4–26.7) | 26.1 (22.8–29.0) | 16.4 (14.5–18.6) | 26.7 (23.4–29.5) | 16.8 (14.9–18.9) |
| 60% efficacy | 771.1 (692.2–857.2) | 219 (182.2–268.3) | 90.5 (57.1–132.2) | 18.7 (8.7–31.5) | 30.7 (27.0–33.8) | 19.2 (16.9–21.6) | 31.3 (27.5–34.4) | 19.6 (17.4–21.9) |
| 70% efficacy | 881.3 (792.1–978.7) | 250.6 (208.8–306.3) | 103.6 (65.1–151.3) | 21.4 (10.0–36) | 35.0 (31.0–38.3) | 21.7 (19.3–24.4) | 35.6 (31.5–38.9) | 22.2 (19.8–24.8) |
| 5 years protection | 474.4 (423.5–530.5) | 131.6 (108.8–162.6) | 56.8 (35.8–83.0) | 11.4 (5.3–19.3) | 16.3 (13.8–18.4) | 9.8 (8.5–11.3) | 17.3 (14.8–19.5) | 10.4 (9.0–11.8) |
| 15 years protection | 740.0 (664.6–822.8) | 212.3 (176.6–259.9) | 86.0 (54.4–125.7) | 18.0 (8.4–30.3) | 31.3 (27.6–34.3) | 20.0 (17.7–22.4) | 31.4 (27.7–34.4) | 20.1 (17.9–22.5) |
| 20 years protection | 790.1 (710.3–877.8) | 227.9 (190–278.7) | 91.4 (57.8–133.6) | 19.2 (9–32.3) | 34.4 (30.5–37.5) | 22.2 (19.8–24.8) | 34.1 (30.2–37.4) | 22.1 (19.9–24.7) |
| Efficacy with current infection at vaccination | 471.4 (402.9–534.9) | 101 (84.2–123.6) | 55.3 (34.1–82.0) | 8.7 (4.1–14.5) | 17.0 (15.7–18.8) | 7.7 (6.9–8.5) | 17.8 (16.4–19.6) | 8.0 (7.2–8.8) |
| Prevention of infection and disease | 816.5 (729.9–914.4) | 238.4 (198.2–293.1) | 95.4 (60.5–140.1) | 20.3 (9.4–34.2) | 33.3 (28.7–37.2) | 21.7 (19.2–24.5) | 33.8 (29.1–37.6) | 22.0 (19.5–24.7) |
| 2036 introduction | 407.7 (363.7–453.5) | 105 (86.8–129.6) | 43.5 (27.4–64.5) | 8.3 (3.9–13.9) | 26.8 (24.4–29.2) | 15.6 (13.8–17.6) | 26.2 (23.6–28.4) | 15.4 (13.8–17.3) |
| Lower coverage | 519.9 (464.8–580.9) | 145.1 (120.1–178.9) | 60.9 (38.5–89.0) | 12.3 (5.7–20.9) | 21.1 (18.4–23.5) | 13.1 (11.5–14.9) | 21.5 (18.7–23.9) | 13.3 (11.7–15.1) |
| Higher coverage | 785.1 (705.6–872.7) | 225.5 (187.7–275.7) | 92.2 (58.2–134.6) | 19.3 (9–32.4) | 31.0 (27.2–34.1) | 19.6 (17.4–22.1) | 31.7 (27.9–34.8) | 20.1 (17.9–22.4) |

| | | | | | | | | |
|---|---|---|---|---|---|---|---|---|
| Older Ages (campaign ages 18–55, routine age 17) | 839.0 (754.7–932.4) | 330.8 (283.5–392.6) | 97.9 (63.2–143.2) | 28.4 (13.4–46.3) | 29.3 (25.1–32.7) | 24.7 (22.6–26.8) | 30.8 (26.4–34.0) | 26 (23.9–28.1) |
| All Adults (campaign ages 19+, routine age 18) | 934.5 (836.3–1,037.4) | 491.8 (433.9–574.9) | 108.1 (70.2–156.6) | 41.9 (20.2–66.2) | 30.5 (25.8–34.4) | 31.8 (30.2–33.7) | 32.2 (27.4–36.1) | 33.9 (32.3–35.8) |
| **BCG-revaccination scenarios** | | | | | | | | |
| Basecase | 358.7 (305.3–402.0) | 112.9 (91.5–142.9) | 44.3 (28.5–64.9) | 10.1 (4.7–17) | 13.3 (9.6–16.2) | 10.1 (8.7–11.8) | 13.7 (10.2–16.4) | 10.1 (8.8–11.8) |
| 70% efficacy | 564.0 (501.0–626.4) | 165.8 (134.8–208.6) | 69.9 (45.3–102.2) | 14.8 (6.8–24.8) | 21.4 (16.6–25.0) | 14.5 (12.6–16.8) | 21.8 (17.3–25.2) | 14.6 (12.8–16.9) |
| 5 years protection | 259.9 (222.0–290.2) | 82.8 (66.7–105.2) | 32.7 (20.9–48.1) | 7.5 (3.4–12.6) | 8.9 (6.3–11.1) | 7.0 (6–8.2) | 9.5 (6.9–11.5) | 7.1 (6.2–8.3) |
| 15 years protection | 407.8 (348.3–457.1) | 128 (103.9–161.7) | 50.0 (32.3–73.6) | 11.3 (5.2–19.1) | 15.8 (11.5–19.0) | 11.8 (10.2–13.8) | 16.0 (12.1–19.0) | 11.7 (10.2–13.7) |
| 20 years protection | 437.8 (374.8–491.2) | 137.2 (111.7–173.2) | 53.5 (34.5–78.5) | 12.1 (5.6–20.4) | 17.4 (12.8–20.8) | 13 (11.2–15.1) | 17.5 (13.2–20.7) | 12.8 (11.2–14.9) |
| Efficacy with any infection at vaccination | 434.3 (389.6–494.2) | 114.1 (92.3–144.6) | 54.0 (34.3–79.9) | 10.2 (4.7–17.1) | 16.3 (13.2–19.1) | 10.1 (8.7–11.9) | 16.7 (13.7–19.3) | 10.2 (8.8–11.9) |
| Prevention of infection and disease | 544.1 (490.0–601.2) | 154.2 (125.2–194.5) | 67.3 (43.3–98.1) | 13.8 (6.4–23.1) | 20.9 (16.9–24.1) | 13.5 (11.7–15.6) | 21.3 (17.5–24.2) | 13.5 (11.8–15.7) |
| 2031 introduction | 237.1 (208.8–263.6) | 66.5 (53.6–84.8) | 28.5 (18.6–41.6) | 5.8 (2.7–9.8) | 11.5 (8.7–13.7) | 7.7 (6.6–9.2) | 12.0 (9.4–14.0) | 7.8 (6.8–9.3) |
| Lower coverage | 322.2 (273.2–361.7) | 102 (82.6–129.3) | 39.7 (25.6–58.3) | 9.1 (4.2–15.3) | 12.0 (8.6–14.7) | 9.2 (7.9–10.8) | 12.4 (9.2–14.8) | 12.6 (10.9–14.4) |
| Higher coverage | 392.3 (336.0–438.9) | 122.9 (99.7–155.4) | 48.5 (31.2–71.1) | 11 (5.1–18.5) | 14.4 (10.5–17.6) | 10.8 (9.4–12.7) | 14.9 (11.2–17.8) | 15 (13.3–16.9) |
| Older Ages (campaign ages 16–34, routine age 15) | 286.9 (195.7–351.7) | 151.6 (124.5–188.2) | 33.2 (19.8–51.4) | 13 (5.9–22) | 10.1 (6.0–13.9) | 12.7 (11–14.6) | 10.1 (6.0–13.9) | 9.2 (8.0–10.8) |
| All Adults (campaign ages 19+, routine age 18) | 223.7 (139.4–286.6) | 184.3 (154.9–222.4) | 25.4 (14.5–40.0) | 15.8 (7.3–26) | 7.9 (4.3–11.4) | 15.1 (13.3–17.0) | 7.9 (4.3–11.3) | 10.9 (9.5–12.7) |

*Cumulative cases and deaths averted between 2025 and 2050 for each of the vaccine scenarios compared to the no-new-vaccine baseline and incidence and mortality rate reductions in 2050 for each of the vaccine scenarios compared to the rate predicted in 2050 with the no-new-vaccine baseline*

#### 10. Economic results

##### 10.1 Delhi Economic Results - M72/AS01E

**Table S10.1 Incremental DALYs averted, incremental costs averted, and ICERs from the health-system and societal perspectives for the M72/AS01E *Vaccine Characteristic and Coverage Scenarios* compared to the no-new-vaccine baseline for Delhi**

| Scenario | Incremental DALYs averted between 2025–2050 (millions) | Health-system perspective | | Societal perspective | |
|---|---|---|---|---|---|
| | | Incremental costs between 2025–2050 ($, millions) | ICERs ($/DALY averted) | Incremental costs between 2025–2050 ($, millions) | ICERs ($/DALY averted) |
| Basecase | 1.5 (1, 2.1) | 5 (-37, 63) | 4 (cost-saving, 47) | -31 (-109, 37) | cost-saving (cost-saving, 26) |
| **Vaccine Characteristic and Coverage Scenarios** | | | | | |
| 60% efficacy | 1.7 (1.2, 2.4) | -15 (-60, 44) | cost-saving (cost-saving, 27) | -59 (-149, 14) | cost-saving (cost-saving, 10) |
| 70% efficacy | 2 (1.3, 2.8) | -34 (-81, 25) | cost-saving (cost-saving, 12) | -85 (-187, -8) | cost-saving |
| 5 years protection | 1.1 (0.7, 1.5) | 34 (-7, 91) | 31 (cost-saving, 89) | 9 (-55, 71) | 8 (cost-saving, 68) |
| 15 years protection | 1.6 (1.1, 2.3) | -8 (-52, 50) | cost-saving (cost-saving, 33) | -49 (-136, 22) | cost-saving (cost-saving, 14) |
| 20 years protection | 1.7 (1.2, 2.4) | -16 (-60, 43) | cost-saving (cost-saving, 26) | -60 (-150, 13) | cost-saving (cost-saving, 9) |
| Prevention of infection and disease | 1.8 (1.2, 2.6) | -22 (-67, 37) | cost-saving (cost-saving, 21) | -68 (-163, 7) | cost-saving (cost-saving, 4) |
| Efficacy with current infection at vaccination | 1.1 (0.7, 1.5) | 36 (-4, 91) | 34 (cost-saving, 100) | 12 (-50, 75) | 11 (cost-saving, 75) |
| 2036 introduction | 0.8 (0.5, 1.1) | 33 (2, 78) | 43 (2, 109) | 16 (-32, 65) | 20 (cost-saving, 89) |
| Lower coverage | 1.2 (0.8, 1.6) | 1 (-32, 45) | 1 (cost-saving, 42) | -28 (-89, 25) | cost-saving (cost-saving, 22) |
| Higher coverage | 1.8 (1.2, 2.5) | 10 (-42, 82) | 6 (cost-saving, 51) | -33 (-128, 50) | cost-saving (cost-saving, 30) |

*Abbreviations: DALYs = disability-adjusted life years, ICERs = incremental cost-effectiveness ratio, US$ = United States Dollar. Values in cells are the mean and 95% uncertainty ranges.*

**Table S10.2 Total vaccination costs, and incremental diagnostic, treatment, and net costs between 2025–2050 for the M72/AS01E scenarios from the health-system perspective for Delhi**

| Scenario | Vaccination costs (US$, millions) | DS-TB diagnostic costs (US$, millions) | RR-TB diagnostic costs (US$, millions) | DS-TB treatment costs (US$, millions) | RR-TB treatment costs (US$, millions) | Incremental cost (US$, millions) |
|---|---|---|---|---|---|---|
| Basecase | 118<br>(80, 173) | -5<br>(-6, -4) | -0.2<br>(-0.8, -0.001) | -75<br>(-91, -60) | -32<br>(-37, -28) | 5<br>(-37, 63) |
| **Policy Scenarios** | | | | | | |
| Older ages (campaign for ages 18–55, routine age 17) | 191<br>(129, 281) | -7<br>(-8, -6) | -0.3<br>(-1.1, -0.001) | -97<br>(-118, -78) | -42<br>(-49, -36) | 45<br>(-21, 139) |
| All adults (campaign for ages 19+, routine age 18) | 235<br>(159, 346) | -8<br>(-9, -6) | -0.3<br>(-1.2, -0.001) | -108<br>(-131, -87) | -46<br>(-54, -40) | 73<br>(-7, 190) |
| **Vaccine Characteristic and Coverage Scenarios** | | | | | | |
| 60% efficacy | 118<br>(80, 173) | -6<br>(-8, -5) | -0.2<br>(-1.0, -0.001) | -88<br>(-107, -71) | -38<br>(-44, -33) | -15<br>(-60, 44) |
| 70% efficacy | 118<br>(80, 173) | -7<br>(-9, -6) | -0.3<br>(-1.1, -0.001) | -101<br>(-123, -81) | -44<br>(-50, -37) | -34<br>(-81, 25) |
| 5 years protection | 118<br>(80, 173) | -4<br>(-5, -3) | -0.1<br>(-0.6, -0.001) | -56<br>(-68, -45) | -24<br>(-28, -21) | 34<br>(-7, 91) |
| 15 years protection | 118<br>(80, 173) | -6<br>(-7, -5) | -0.2<br>(-0.9, -0.001) | -84<br>(-102, -67) | -36<br>(-42, -31) | -8<br>(-52, 50) |
| 20 years protection | 118<br>(80, 173) | -6<br>(-8, -5) | -0.2<br>(-1.0, -0.001) | -89<br>(-108, -71) | -38<br>(-44, -33) | -16<br>(-60, 43) |
| Prevention of infection and disease | 118<br>(80, 173) | -7<br>(-8, -5) | -0.2<br>(-1.0, -0.001) | -93<br>(-113, -74) | -40<br>(-47, -34) | -22<br>(-67, 37) |
| Efficacy with current infection at vaccination | 118<br>(80, 173) | -4<br>(-5, -3) | -0.1<br>(-0.6, -0.001) | -54<br>(-66, -44) | -23<br>(-27, -20) | 36<br>(-4, 91) |
| 2036 introduction | 93<br>(63, 137) | -3<br>(-3, -2) | -0.1<br>(-0.4, -0.001) | -40<br>(-49, -32) | -17<br>(-20, -15) | 33<br>(2, 78) |
| Lower coverage | 90<br>(61, 132) | -4<br>(-5, -3) | -0.2<br>(-0.7, -0.001) | -59<br>(-72, -47) | -26<br>(-30, -22) | 1<br>(-32, 45) |
| Higher coverage | 146<br>(99, 214) | -6<br>(-8, -5) | -0.2<br>(-1.0, -0.001) | -90<br>(-110, -72) | -39<br>(-45, -33) | 10<br>(-42, 82) |

*Abbreviations: DS-TB = drug-susceptible tuberculosis, RR-TB = rifampicin resistant tuberculosis, US$ = United States Dollars. Values in cells are the mean and 95% uncertainty ranges.*

**Table S10.3 Total vaccination costs, and incremental diagnostic, treatment, and net costs between 2025–2050 for the M72/AS01E scenarios from the societal perspective for Delhi**

| Scenario | Vaccination costs (US$, millions) | Diagnostic costs (DS + RR-TB) (US$, millions) | Treatment costs (DS + RR-TB) (US$, millions) | Non-medical costs (US$, millions) | Indirect costs (US$, millions) | Incremental cost (US$, millions) |
|---|---|---|---|---|---|---|
| Basecase | 124 (84, 180) | -5 (-7, -4) | -107 (-128, -89) | -13 (-22, -7) | -29 (-99, -1) | -31 (-109, 37) |
| **Policy Scenarios** | | | | | | |
| Older ages (campaign for ages 18–55, routine age 17) | 202 (137, 292) | -7 (-9, -6) | -139 (-166, -114) | -17 (-28, -9) | -38 (-128, -2) | 1 (-110, 106) |
| All adults (campaign for ages 19+, routine age 18) | 248 (168, 359) | -8 (-10, -6) | -154 (-184, -127) | -19 (-32, -10) | -42 (-140, -2) | 25 (-101, 153) |
| **Vaccine Characteristic and Coverage Scenarios** | | | | | | |
| 60% efficacy | 124 (84, 180) | -6 (-8, -5) | -126 (-151, -104) | -16 (-26, -8) | -35 (-117, -2) | -59 (-149, 14) |
| 70% efficacy | 124 (84, 180) | -7 (-9, -6) | -144 (-172, -119) | -18 (-29, -10) | -40 (-134, -2) | -85 (-187, -8) |
| 5 years protection | 124 (84, 180) | -4 (-5, -3) | -80 (-95, -66) | -10 (-16, -5) | -22 (-74, -1) | 9 (-55, 71) |
| 15 years protection | 124 (84, 180) | -6 (-7, -5) | -120 (-143, -99) | -15 (-24, -8) | -33 (-111, -1) | -49 (-136, 22) |
| 20 years protection | 124 (84, 180) | -7 (-8, -5) | -127 (-151, -105) | -16 (-26, -8) | -35 (-118, -2) | -60 (-150, 13) |
| Prevention of infection and disease | 124 (84, 180) | -7 (-8, -6) | -133 (-159, -109) | -17 (-27, -9) | -36 (-123, -2) | -68 (-163, 7) |
| Efficacy with current infection at vaccination | 124 (84, 179) | -4 (-5, -3) | -78 (-93, -64) | -10 (-16, -5) | -21 (-72, -1) | 12 (-50, 75) |
| 2036 introduction | 98 (67, 142) | -3 (-4, -2) | -57 (-68, -47) | -7 (-12, -4) | -16 (-53, -1) | 16 (-32, 65) |
| Lower coverage | 95 (65, 137) | -4 (-5, -4) | -85 (-101, -70) | -11 (-17, -6) | -23 (-78, -1) | -28 (-89, 25) |
| Higher coverage | 154 (104, 222) | -7 (-8, -5) | -129 (-154, -106) | -16 (-26, -9) | -35 (-119, -2) | -33 (-128, 50) |

*Abbreviations: DS-TB = drug-susceptible tuberculosis, RR-TB = rifampicin resistant tuberculosis, US$ = United States Dollars. Values in cells are the mean and 95% uncertainty ranges.*

#### 10.2 Delhi Economic Results - BCG-revaccination

**Table S10.4 Incremental DALYs averted, incremental costs averted, and ICERs from the health-system and societal perspectives for the BCG-revaccination *Vaccine Characteristic and Coverage Scenarios* compared to the no-new-vaccine baseline for Delhi**

| Scenario | Incremental DALYs averted between 2025–2050 (millions) | Health-system perspective | | Societal perspective | |
|---|---|---|---|---|---|
| | | Incremental costs between 2025–2050 ($, thousands) | ICERs ($/DALY averted) | Incremental costs between 2025–2050 ($, millions) | ICERs ($/DALY averted) |
| Basecase | 0.9 (0.6, 1.3) | -38 (-58, -13) | cost-saving | -59 (-103, -26) | cost-saving |
| **Vaccine Characteristic and Coverage Scenarios** | | | | | |
| 70% efficacy | 1.5 (1.0, 2.0) | -74 (-100, -46) | cost-saving | -110 (-176, -67) | cost-saving |
| 5 years protection | 0.7 (0.5, 1.0) | -21 (-39, 3) | cost-saving (cost-saving, 5) | -36 (-69, -8) | cost-saving |
| 15 years protection | 1.1 (0.7, 1.5) | -46 (-68, -20) | cost-saving | -71 (-120, -35) | cost-saving |
| 20 years protection | 1.1 (0.8, 1.6) | -51 (-74, -25) | cost-saving | -78 (-129, -41) | cost-saving |
| Prevention of infection and disease | 1.4 (1.0, 1.9) | -71 (-96, -43) | cost-saving | -105 (-168, -64) | cost-saving |
| Efficacy with any infection at vaccination | 1.1 (0.8, 1.6) | -52 (-74, -25) | cost-saving | -79 (-130, -43) | cost-saving |
| 2031 introduction | 0.6 (0.4, 0.8) | -20 (-33, -4) | cost-saving | -33 (-59, -12) | cost-saving |
| Lower coverage | 0.8 (0.6, 1.2) | -34 (-52, -12) | cost-saving | -54 (-93, -24) | cost-saving |
| Higher coverage | 1.0 (0.7, 1.4) | -41 (-63, -13) | cost-saving | -65 (-112, -28) | cost-saving |

*Abbreviations: DALYs = disability-adjusted life years, ICERs = incremental cost-effectiveness ratio, US$ = United States Dollar. Values in cells are the mean and 95% uncertainty ranges.*

**Table 10.5 Total vaccination costs, and incremental diagnostic, treatment, and net costs between 2025–2050 for the BCG-revaccination scenarios from the health-system perspective for Delhi**

| Scenario | Vaccination costs (US$, millions) | DS-TB diagnostic costs (US$, millions) | RR-TB diagnostic costs (US$, millions) | DS-TB treatment costs (US$, millions) | RR-TB treatment costs (US$, millions) | Incremental cost (US$, millions) |
|---|---|---|---|---|---|---|
| Basecase | 27<br>(12, 49) | -3<br>(-4, -2) | -0.1<br>(-0.5, -0.001) | -43<br>(-53, -34) | -18<br>(-22, -15) | -38<br>(-58, -13) |
| **Policy Scenarios** | | | | | | |
| Older ages (campaign for ages 16-34, routine age 15) | 48<br>(20, 88) | -2<br>(-3, -2) | -0.09<br>(-0.4, 0) | -35<br>(-46, -23) | -15<br>(-19, -10) | -4<br>(-36, 40) |
| All adults (campaign for ages 19+, routine age 18) | 95<br>(40, 176) | -2<br>(-3, -1) | -0.07<br>(-0.3, 0) | -27<br>(-37, -16) | -12<br>(-16, -7) | 55<br>(-3, 139) |
| **Vaccine Characteristic and Coverage Scenarios** | | | | | | |
| 70% efficacy | 27<br>(12, 49) | -5<br>(-6, -4) | -0.2<br>(-0.7, -0.001) | -67<br>(-83, -54) | -29<br>(-34, -24) | -74<br>(-100, -46) |
| 5 years protection | 27<br>(12, 49) | -2<br>(-3, -2) | -0.08<br>(-0.3, 0) | -32<br>(-39, -25) | -14<br>(-16, -11) | -21<br>(-39, 3) |
| 15 years protection | 27<br>(12, 49) | -3<br>(-4, -3) | -0.1<br>(-0.5, -0.001) | -48<br>(-60, -38) | -21<br>(-25, -17) | -46<br>(-68, -20) |
| 20 years protection | 27<br>(12, 49) | -4<br>(-4, -3) | -0.1<br>(-0.6, -0.001) | -52<br>(-64, -41) | -22<br>(-26, -18) | -51<br>(-74, -25) |
| Prevention of infection and disease | 27<br>(12, 49) | -5<br>(-6, -4) | -0.2<br>(-0.7, -0.001) | -65<br>(-80, -52) | -28<br>(-33, -24) | -71<br>(-96, -43) |
| Efficacy with any infection at vaccination | 27<br>(12, 50) | -4<br>(-5, -3) | -0.1<br>(-0.6, -0.001) | -52<br>(-65, -42) | -23<br>(-27, -19) | -52<br>(-74, -25) |
| 2031 introduction | 18<br>(8, 33) | -2<br>(-2, -1) | -0.07<br>(-0.3, 0) | -25<br>(-31, -20) | -11<br>(-13, -9) | -20<br>(-33, -4) |
| Lower coverage | 24<br>(10, 43) | -3<br>(-3, -2) | -0.1<br>(-0.4, 0) | -38<br>(-48, -30) | -17<br>(-20, -14) | -34<br>(-52, -12) |
| Higher coverage | 30<br>(13, 55) | -3<br>(-4, -3) | -0.1<br>(-0.5, -0.001) | -47<br>(-58, -37) | -20<br>(-24, -17) | -41<br>(-63, -13) |

*Abbreviations: DS-TB = drug-susceptible tuberculosis, RR-TB = rifampicin resistant tuberculosis, US$ = United States Dollars. Values in cells are the mean and 95% uncertainty ranges.*

**Table S10.6 Total vaccination costs, and incremental diagnostic, treatment, and net costs between 2025–2050 for the BCG-revaccination scenarios from the societal perspective for Delhi**

| Scenario | Vaccination costs (US$, millions) | Diagnostic costs (DS + RR-TB) (US$, millions) | Treatment costs (DS + RR-TB) (US$, millions) | Non-medical costs (US$, millions) | Indirect costs (US$, millions) | Incremental cost (US$, millions) |
|---|---|---|---|---|---|---|
| Basecase | 30<br>(14, 51) | -3<br>(-4, -2) | -61<br>(-74, -49) | -8<br>(-13, -4) | -17<br>(-55, -1) | -59<br>(-103, -26) |
| **Policy Scenarios** | | | | | | |
| Older ages (campaign for ages 16-34, routine age 15) | 53<br>(24, 93) | -3<br>(-3, -2) | -50<br>(-65, -33) | -6<br>(-11, -3) | -14<br>(-44, -1) | -19<br>(-65, 29) |
| All adults (campaign for ages 19+, routine age 18) | 105<br>(46, 185) | -2<br>(-3, -1) | -38<br>(-53, -24) | -5<br>(-9, -2) | -10<br>(-34, 0) | 49<br>(-14, 136) |
| **Vaccine Characteristic and Coverage Scenarios** | | | | | | |
| 70% efficacy | 30<br>(14, 51) | -5<br>(-6, -4) | -97<br>(-116, -79) | -12<br>(-20, -7) | -26<br>(-88, -1) | -110<br>(-176, -67) |
| 5 years protection | 30<br>(14, 51) | -2<br>(-3, -2) | -45<br>(-55, -36) | -6<br>(-9, -3) | -12<br>(-41, -1) | -36<br>(-69, -8) |
| 15 years protection | 29<br>(14, 51) | -4<br>(-4, -3) | -69<br>(-84, -55) | -9<br>(-14, -5) | -19<br>(-62, -1) | -71<br>(-120, -35) |
| 20 years protection | 29<br>(14, 51) | -4<br>(-5, -3) | -74<br>(-89, -59) | -9<br>(-15, -5) | -20<br>(-67, -1) | -78<br>(-129, -41) |
| Prevention of infection and disease | 30<br>(14, 51) | -5<br>(-6, -4) | -93<br>(-111, -77) | -12<br>(-19, -6) | -26<br>(-85, -1) | -105<br>(-168, -64) |
| Efficacy with any infection at vaccination | 30<br>(14, 52) | -4<br>(-5, -3) | -75<br>(-91, -61) | -9<br>(-15, -5) | -21<br>(-68, -1) | -79<br>(-130, -43) |
| 2031 introduction | 20<br>(9, 34) | -2<br>(-2, -1) | -36<br>(-44, -29) | -5<br>(-7, -2) | -10<br>(-33, 0) | -33<br>(-59, -12) |
| Lower coverage | 26<br>(12, 45) | -3<br>(-3, -2) | -55<br>(-67, -44) | -7<br>(-11, -4) | -15<br>(-49, -1) | -54<br>(-93, -24) |
| Higher coverage | 33<br>(15, 57) | -3<br>(-4, -3) | -67<br>(-81, -54) | -8<br>(-14, -5) | -18<br>(-60, -1) | -65<br>(-112, -28) |

*Abbreviations: DS-TB = drug-susceptible tuberculosis, RR-TB = rifampicin resistant tuberculosis, US$ = United States Dollars. Values in cells are the mean and 95% uncertainty ranges.*

#### 10.3 Gujarat Economic Results - M72/AS01$_E$

**Table S10.7 Incremental DALYs averted, incremental costs averted, and ICERs from the health-system and societal perspectives for the M72/AS01$_E$ *Vaccine Characteristic and Coverage Scenarios* compared to the no-new-vaccine baseline for Gujarat**

| Scenario | Incremental DALYs averted between 2025–2050 (millions) | Health-system perspective | | Societal perspective | |
|---|---|---|---|---|---|
| | | Incremental costs between 2025–2050 ($, millions) | ICERs ($/DALY averted) | Incremental costs between 2025–2050 ($, millions) | ICERs ($/DALY averted) |
| Basecase | 0.3<br>(0.2, 0.5) | 332<br>(213, 505) | 1 078<br>(567, 2 402) | 385<br>(251, 556) | 1 250<br>(663, 2 649) |
| **Vaccine Characteristic and Coverage Scenarios** | | | | | |
| 60% efficacy | 0.4<br>(0.2, 0.6) | 327<br>(208, 500) | 898<br>(469, 2 007) | 377<br>(242, 549) | 1 037<br>(545, 2 212) |
| 70% efficacy | 0.4<br>(0.2, 0.7) | 321<br>(203, 494) | 770<br>(397, 1 725) | 369<br>(234, 539) | 884<br>(464, 1 901) |
| 5 years protection | 0.2<br>(0.1, 0.4) | 341<br>(222, 513) | 1 511<br>(805, 3 336) | 398<br>(263, 569) | 1 762<br>(960, 3 730) |
| 15 years protection | 0.3<br>(0.2, 0.5) | 328<br>(210, 501) | 947<br>(496, 2 106) | 379<br>(245, 552) | 1 094<br>(576, 2 325) |
| 20 years protection | 0.4<br>(0.2, 0.6) | 326<br>(208, 499) | 882<br>(459, 1 960) | 376<br>(241, 547) | 1 017<br>(538, 2 169) |
| Prevention of infection and disease | 0.4<br>(0.2, 0.6) | 323<br>(206, 496) | 818<br>(425, 1 852) | 372<br>(238, 543) | 942<br>(491, 2 020) |
| Efficacy with current infection at vaccination | 0.2<br>(0.1, 0.3) | 347<br>(228, 518) | 2052<br>(1 104, 4 466) | 406<br>(271, 578) | 2402<br>(1313, 4 994) |
| 2036 introduction | 0.1<br>(0.1, 0.2) | 260<br>(170, 390) | 1 737<br>(928, 3 808) | 304<br>(201, 433) | 2029<br>(1100, 4 349) |
| Lower coverage | 0.2<br>(0.1, 0.4) | 256<br>(164, 389) | 1 067<br>(561, 2 395) | 297<br>(194, 428) | 1236<br>(651, 2 633) |
| Higher coverage | 0.4<br>(0.2, 0.6) | 409<br>(262, 621) | 1 095<br>(576, 2 424) | 475<br>(310, 685) | 1269<br>(679, 2 688) |

*Abbreviations: DALYs = disability-adjusted life years, ICERs = incremental cost-effectiveness ratio, US$ = United States Dollar. Values in cells are the mean and 95% uncertainty ranges.*

**Table S10.8** **Total vaccination costs, and incremental diagnostic, treatment, and net costs between 2025–2050 for the M72/AS01$_E$ scenarios from the health-system perspective for Gujarat**

| Scenario | Vaccination costs (US$, millions) | DS-TB diagnostic costs (US$, millions) | RR-TB diagnostic costs (US$, millions) | DS-TB treatment costs (US$, millions) | RR-TB treatment costs (US$, millions) | Incremental cost (US$, millions) |
|---|---|---|---|---|---|---|
| Basecase | 366 (248, 536) | -2 (-2, -1) | -0.036 (-0.157, 0) | -25 (-33, -19) | -6 (-8, -5) | 332 (213, 505) |
| **Policy Scenarios** | | | | | | |
| Older ages (campaign for ages 18–55, routine age 17) | 573 (388, 841) | -3 (-4, -2) | -0.066 (-0.282, 0) | -46 (-59, -35) | -11 (-13, -9) | 513 (327, 784) |
| All adults (campaign for ages 19+, routine age 18) | 713 (482, 1049) | -5 (-6, -4) | -0.098 (-0.41, 0) | -68 (-86, -53) | -16 (-20, -13) | 624 (393, 960) |
| **Vaccine Characteristic and Coverage Scenarios** | | | | | | |
| 60% efficacy | 366 (248, 536) | -2 (-3, -2) | -0.043 (-0.185, 0) | -30 (-39, -22) | -7 (-9, -6) | 327 (208, 500) |
| 70% efficacy | 366 (248, 536) | -2 (-3, -2) | -0.049 (-0.212, 0) | -34 (-45, -26) | -8 (-10, -6) | 321 (203, 494) |
| 5 years protection | 365 (248, 536) | -1 (-2, -1) | -0.026 (-0.114, 0) | -18 (-24, -14) | -4 (-6, -3) | 341 (222, 513) |
| 15 years protection | 366 (248, 536) | -2 (-3, -2) | -0.041 (-0.177, 0) | -28 (-37, -21) | -7 (-9, -5) | 328 (210, 501) |
| 20 years protection | 366 (248, 536) | -2 (-3, -2) | -0.043 (-0.189, 0) | -30 (-40, -23) | -7 (-9, -6) | 326 (208, 499) |
| Prevention of infection and disease | 366 (248, 536) | -2 (-3, -2) | -0.046 (-0.198, 0) | -32 (-42, -24) | -8 (-10, -6) | 323 (206, 496) |
| Efficacy with current infection at vaccination | 365 (248, 536) | -1 (-1, -1) | -0.02 (-0.087, 0) | -14 (-18, -11) | -3 (-4, -3) | 347 (228, 518) |
| 2036 introduction | 276 (187, 405) | -1 (-1, -1) | -0.018 (-0.076, 0) | -12 (-16, -9) | -3 (-4, -2) | 260 (170, 390) |
| Lower coverage | 282 (191, 413) | -1 (-2, -1) | -0.028 (-0.121, 0) | -20 (-26, -15) | -5 (-6, -4) | 256 (164, 389) |
| Higher coverage | 449 (304, 660) | -2 (-3, -2) | -0.044 (-0.191, 0) | -31 (-40, -23) | -7 (-9, -6) | 409 (262, 621) |

*Abbreviations: DS-TB = drug-susceptible tuberculosis, RR-TB = rifampicin resistant tuberculosis, US$ = United States Dollars. Values in cells are the mean and 95% uncertainty ranges.*

**Table S10.9 Total vaccination costs, and incremental diagnostic, treatment, and net costs between 2025–2050 for the M72/AS01$_E$ scenarios from the societal perspective for Gujarat**

| Scenario | Vaccination costs (US$, millions) | Diagnostic costs (DS + RR-TB) (US$, millions) | Treatment costs (DS + RR-TB) (US$, millions) | Non-medical costs (US$, millions) | Indirect costs (US$, millions) | Incremental cost (US$, millions) |
|---|---|---|---|---|---|---|
| Basecase | 432 (298, 605) | -2 (-2, -1) | -31 (-41, -24) | -4 (-7, -2) | -10 (-33, 0) | 385 (251, 556) |
| **Policy Scenarios** | | | | | | |
| Older ages (campaign for ages 18–55, routine age 17) | 678 (467, 949) | -3 (-4, -3) | -57 (-72, -44) | -8 (-13, -4) | -18 (-59, -1) | 593 (382, 863) |
| All adults (campaign for ages 19+, routine age 18) | 845 (582, 1183) | -5 (-6, -4) | -85 (-106, -67) | -12 (-20, -6) | -26 (-87, -1) | 717 (452, 1055) |
| **Vaccine Characteristic and Coverage Scenarios** | | | | | | |
| 60% efficacy | 432 (298, 605) | -2 (-3, -2) | -37 (-48, -28) | -5 (-9, -3) | -11 (-39, 0) | 377 (242, 549) |
| 70% efficacy | 432 (298, 605) | -2 (-3, -2) | -42 (-55, -32) | -6 (-10, -3) | -13 (-44, -1) | 369 (234, 539) |
| 5 years protection | 432 (298, 605) | -1 (-2, -1) | -23 (-30, -17) | -3 (-5, -2) | -7 (-24, 0) | 398 (263, 569) |
| 15 years protection | 432 (298, 605) | -2 (-3, -2) | -35 (-46, -27) | -5 (-8, -3) | -11 (-37, 0) | 379 (245, 552) |
| 20 years protection | 432 (298, 605) | -2 (-3, -2) | -38 (-49, -29) | -5 (-9, -3) | -12 (-39, 0) | 376 (241, 547) |
| Prevention of infection and disease | 432 (298, 605) | -2 (-3, -2) | -40 (-52, -30) | -6 (-9, -3) | -12 (-42, -1) | 372 (238, 543) |
| Efficacy with current infection at vaccination | 432 (298, 605) | -1 (-1, -1) | -17 (-23, -13) | -2 (-4, -1) | -5 (-18, 0) | 406 (271, 578) |
| 2036 introduction | 327 (225, 457) | -1 (-1, -1) | -15 (-20, -12) | -2 (-4, -1) | -5 (-16, 0) | 304 (201, 433) |
| Lower coverage | 333 (229, 466) | -1 (-2, -1) | -24 (-32, -18) | -3 (-6, -2) | -8 (-26, 0) | 297 (194, 428) |
| Higher coverage | 532 (366, 744) | -2 (-3, -2) | -38 (-49, -29) | -5 (-9, -3) | -12 (-40, 0) | 475 (310, 685) |

*Abbreviations: DS-TB = drug-susceptible tuberculosis, RR-TB = rifampicin resistant tuberculosis, US$ = United States Dollars. Values in cells are the mean and 95% uncertainty ranges.*

#### 10.4 Gujarat Economic Results - BCG-revaccination

**Table S10.10 Incremental DALYs averted, incremental costs averted, and ICERs from the health-system and societal perspectives for the BCG-revaccination *Vaccine Characteristic and Coverage Scenarios* compared to the no-new-vaccine baseline for Gujarat**

| Scenario | Incremental DALYs averted between 2025–2050 (millions) | Health-system perspective | | Societal perspective | |
|---|---|---|---|---|---|
| | | Incremental costs between 2025–2050 ($, millions) | ICERs ($/DALY averted) | Incremental costs between 2025–2050 ($, millions) | ICERs ($/DALY averted) |
| Basecase | 0.2 (0.1, 0.3) | 77 (21, 158) | 351 (91, 973) | 99 (35, 177) | 452 (143, 1 139) |
| **Vaccine Characteristic and Coverage Scenarios** | | | | | |
| 70% efficacy | 0.3 (0.2, 0.5) | 67 (12, 148) | 208 (33, 609) | 85 (19, 164) | 263 (58, 709) |
| 5 years protection | 0.2 (0.1, 0.3) | 82 (26, 162) | 497 (149, 1329) | 106 (43, 186) | 644 (235, 1 582) |
| 15 years protection | 0.2 (0.1, 0.4) | 74 (19, 155) | 303 (71, 853) | 95 (31, 174) | 389 (112, 995) |
| 20 years protection | 0.3 (0.1, 0.4) | 72 (17, 153) | 279 (61, 789) | 93 (28, 172) | 357 (98, 919) |
| Prevention of infection and disease | 0.3 (0.2, 0.5) | 69 (14, 150) | 230 (40, 665) | 88 (22, 167) | 292 (72, 772) |
| Efficacy with any infection at vaccination | 0.2 (0.1, 0.4) | 76 (21, 157) | 346 (89, 964) | 99 (34, 177) | 446 (140, 1 127) |
| 2031 introduction | 0.1 (0.1, 0.2) | 52 (16, 105) | 454 (132, 1255) | 68 (26, 120) | 589 (206, 1 466) |
| Lower coverage | 0.2 (0.1, 0.3) | 67 (19, 138) | 343 (90, 950) | 87 (30, 155) | 440 (139, 1 115) |
| Higher coverage | 0.2 (0.1, 0.4) | 86 (24, 176) | 360 (94, 994) | 111 (40, 199) | 465 (150, 1 162) |

*Abbreviations: DALYs = disability-adjusted life years, ICERs = incremental cost-effectiveness ratio, US$ = United States Dollar. Values in cells are the mean and 95% uncertainty ranges.*

**Table S10.11** **Total vaccination costs, and incremental diagnostic, treatment, and net costs between 2025–2050 for the BCG-revaccination scenarios from the health-system perspective for Gujarat**

| Scenario | Vaccination costs (US$, millions) | DS-TB diagnostic costs (US$, millions) | RR-TB diagnostic costs (US$, millions) | DS-TB treatment costs (US$, millions) | RR-TB treatment costs (US$, millions) | Incremental cost (US$, millions) |
|---|---|---|---|---|---|---|
| Basecase | 97 (42, 178) | -1 (-2, -1) | -0.02 (-0.1, 0) | -16 (-21, -12) | -4 (-5, -3) | 77 (21, 158) |
| **Policy Scenarios** | | | | | | |
| Older ages (campaign for ages 16-34, routine age 15) | 152 (64, 279) | -2 (-2, -1) | -0.03 (-0.1, 0) | -22 (-29, -16) | -5 (-7, -4) | 124 (35, 252) |
| All adults (campaign for ages 19+, routine age 18) | 294 (123, 543) | -2 (-2, -1) | -0.04 (-0.2, 0) | -26 (-34, -20) | -6 (-8, -5) | 260 (86, 510) |
| **Vaccine Characteristic and Coverage Scenarios** | | | | | | |
| 70% efficacy | 97 (42, 178) | -2 (-2, -1) | -0.03 (-0.1, 0) | -23 (-31, -17) | -6 (-7, -4) | 67 (12, 148) |
| 5 years protection | 97 (42, 178) | -1 (-1, -1) | -0.02 (-0.07, 0) | -12 (-16, -9) | -3 (-4, -2) | 82 (26, 162) |
| 15 years protection | 97 (42, 178) | -1 (-2, -1) | -0.03 (-0.1, 0) | -18 (-24, -13) | -4 (-6, -3) | 74 (19, 155) |
| 20 years protection | 97 (42, 178) | -1 (-2, -1) | -0.03 (-0.1, 0) | -19 (-26, -14) | -5 (-6, -3) | 72 (17, 153) |
| Prevention of infection and disease | 97 (42, 178) | -2 (-2, -1) | -0.03 (-0.1, 0) | -22 (-29, -16) | -5 (-7, -4) | 69 (14, 150) |
| Efficacy with any infection at vaccination | 98 (42, 178) | -1 (-2, -1) | -0.02 (-0.1, 0) | -16 (-22, -12) | -4 (-5, -3) | 76 (21, 157) |
| 2031 introduction | 63 (27, 116) | -1 (-1, 0) | -0.01 (-0.05, 0) | -8 (-11, -6) | -2 (-3, -2) | 52 (16, 105) |
| Lower coverage | 86 (37, 157) | -1 (-1, -1) | -0.02 (-0.09, 0) | -14 (-19, -11) | -3 (-4, -3) | 67 (19, 138) |
| Higher coverage | 109 (46, 198) | -1 (-2, -1) | -0.03 (-0.1, 0) | -17 (-23, -13) | -4 (-5, -3) | 86 (24, 176) |

*Abbreviations: DS-TB = drug-susceptible tuberculosis, RR-TB = rifampicin resistant tuberculosis, US$ = United States Dollars. Values in cells are the mean and 95% uncertainty ranges.*

**Table S10.12 Total vaccination costs, and incremental diagnostic, treatment, and net costs between 2025–2050 for the BCG-revaccination scenarios from the societal perspective for Gujarat**

| Scenario | Vaccination costs (US$, millions) | Diagnostic costs (DS + RR-TB) (US$, millions) | Treatment costs (DS + RR-TB) (US$, millions) | Non-medical costs (US$, millions) | Indirect costs (US$, millions) | Incremental cost (US$, millions) |
|---|---|---|---|---|---|---|
| Basecase | 128 (64, 208) | -1 (-2, -1) | -20 (-26, -15) | -3 (-5, -1) | -6 (-21, 0) | 99 (35, 177) |
| **Policy Scenarios** | | | | | | |
| Older ages (campaign for ages 16-34, routine age 15) | 202 (101, 328) | -2 (-2, -1) | -27 (-35, -20) | -4 (-6, -2) | -8 (-28, 0) | 161 (61, 288) |
| All adults (campaign for ages 19+, routine age 18) | 392 (195, 643) | -2 (-2, -1) | -33 (-42, -25) | -5 (-8, -2) | -10 (-34, 0) | 342 (144, 591) |
| **Vaccine Characteristic and Coverage Scenarios** | | | | | | |
| 70% efficacy | 128 (64, 208) | -2 (-2, -1) | -29 (-38, -22) | -4 (-7, -2) | -9 (-31, 0) | 85 (19, 164) |
| 5 years protection | 128 (64, 208) | -1 (-1, -1) | -15 (-20, -11) | -2 (-3, -1) | -5 (-15, 0) | 106 (43, 186) |
| 15 years protection | 128 (64, 208) | -1 (-2, -1) | -22 (-29, -17) | -3 (-5, -2) | -7 (-23, 0) | 95 (31, 174) |
| 20 years protection | 128 (64, 208) | -1 (-2, -1) | -24 (-31, -18) | -3 (-6, -2) | -7 (-25, 0) | 93 (28, 172) |
| Prevention of infection and disease | 128 (64, 208) | -2 (-2, -1) | -27 (-36, -20) | -4 (-6, -2) | -8 (-29, 0) | 88 (22, 167) |
| Efficacy with any infection at vaccination | 128 (64, 208) | -1 (-2, -1) | -20 (-27, -15) | -3 (-5, -1) | -6 (-21, 0) | 99 (34, 177) |
| 2031 introduction | 84 (42, 136) | -1 (-1, 0) | -10 (-14, -8) | -1 (-2, -1) | -3 (-11, 0) | 68 (26, 120) |
| Lower coverage | 113 (57, 183) | -1 (-1, -1) | -18 (-24, -13) | -2 (-4, -1) | -6 (-19, 0) | 87 (30, 155) |
| Higher coverage | 143 (72, 233) | -1 (-2, -1) | -21 (-29, -16) | -3 (-5, -2) | -7 (-23, 0) | 111 (40, 199) |

*Abbreviations: DS-TB = drug-susceptible tuberculosis, RR-TB = rifampicin resistant tuberculosis, US$ = United States Dollars. Values in cells are the mean and 95% uncertainty ranges.*
